# Reduced Risk of Liver Related Events Among Patients Receiving Individualized Nutrition-Focused Remote Care in the United States

**DOI:** 10.1101/2025.10.24.25338753

**Authors:** Shaminie J Athinarayanan, Adam J Wolfberg, Priya V Shanmugam, Bilal A Hameed, Maurizio Bonacini

**Author notes:** **Address for correspondence:** Shaminie J Athinarayanan, Virta Health, 3513 Brighton Blvd, Suite 310, Denver, CO 80216, USA.

## Abstract

**Background and Aims:** Metabolic dysfunction–associated steatotic liver disease (MASLD) and its progressive form, metabolic dysfunction–associated steatohepatitis (MASH), lead to significant morbidity and mortality in adults with type 2 diabetes (T2D) and obesity. This study evaluated whether participation in an individualized, nutrition-focused telemedicine care model emphasizing carbohydrate reduction (Virta Individualized Nutrition Therapy, VINT) was associated with reduced onset of MASLD, MASH, and advanced liver disease.

**Approach and Results:** Adults with T2D, prediabetes, overweight, or obesity who enrolled in VINT (2015–2024) were identified in the Komodo Healthcare Map and matched 1:1 to usual care (UC) controls (n = 5,031 per group). Using three complementary analytic approaches, incidence and time-to-event analyses were performed for new-onset liver disease. Across all strategies, VINT participants consistently showed lower incidence of any liver-related diagnosis (27.8 vs 42.8 per 1,000 person-years; HR = 0.61, p < 0.001), MASH and beyond (4.2 vs 10.7; HR = 0.38, p < 0.001), advanced liver disease (2.8 vs 8.7; HR = 0.33, p < 0.001) and any liver complications (2.0 vs 7.7; HR = 0.25, p < 0.001). VINT participants who lost ≥15% body weight was at lower risk of new-onset liver disease (21.2 vs 31.8 per 1,000 person-years; HR = 0.66, p = 0.02) compared to VINT participants who lost less weight.

**Conclusions:** Participation in individualized nutrition-focused telemedicine care was associated with significantly lower incidence and risk of new-onset MASLD, MASH, and advanced liver disease. These findings support lifestyle-first interventions that is potentially scalable to reduce liver disease burden in adults with T2D and obesity.

**Infographic:** 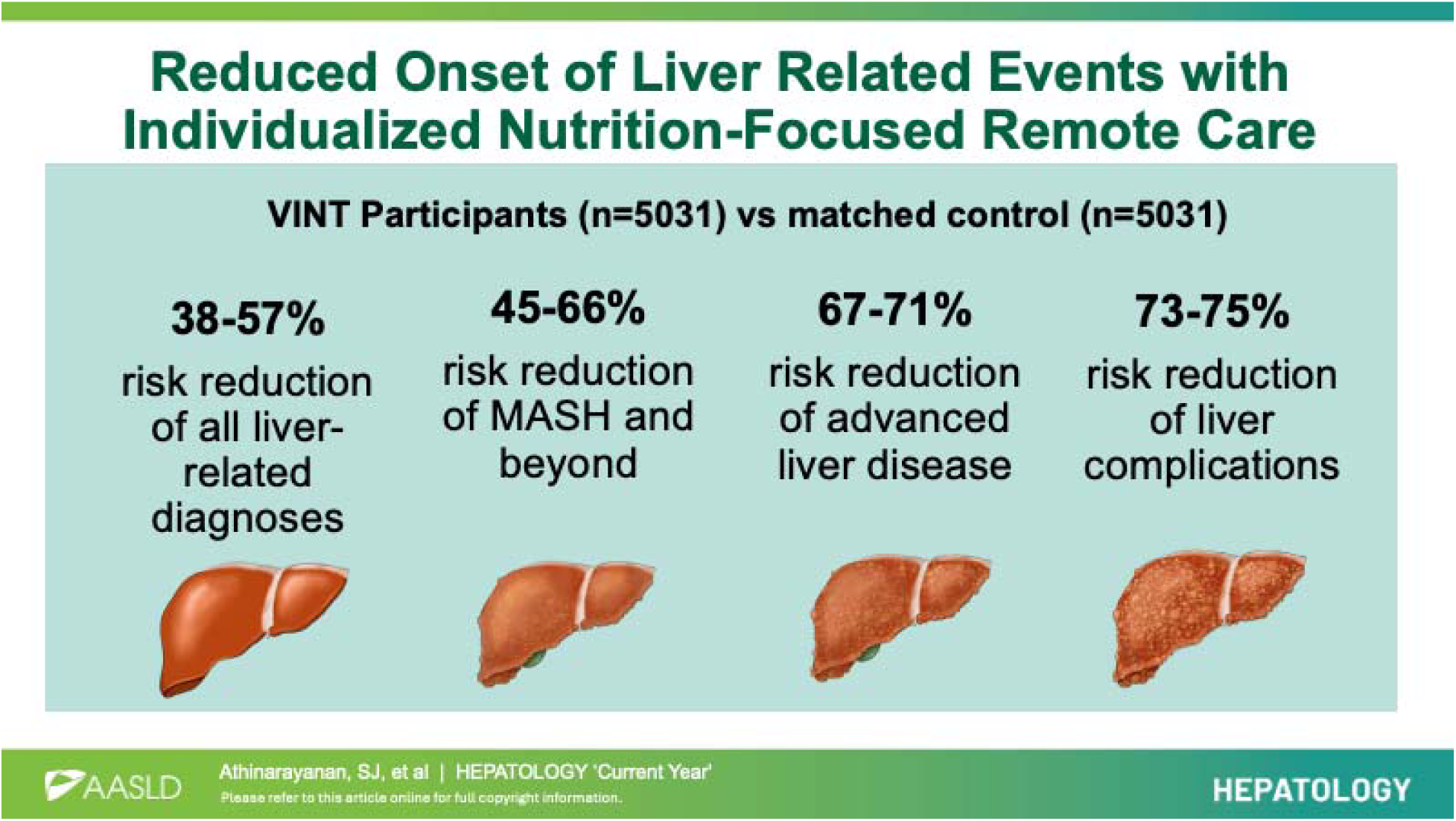

**Plain Language Summary:** People with type 2 diabetes and obesity are more likely to develop liver conditions such as metabolic dysfunction–associated steatotic liver disease (MASLD) and metabolic dysfunction–associated steatohepatitis (MASH), which can progress to cirrhosis, decompensation or liver cancer. This study examined whether a nutrition-focused remote care program could help prevent these liver diseases. The Virta Individualized Nutrition Therapy (VINT) program uses telemedicine and personalized, carbohydrate-reduced nutrition to support long-term weight loss and metabolic health. Using health claims data, researchers compared more than 5,000 participants in the VINT program with matched individuals receiving standard of care. Those in the VINT group had significantly fewer new cases of MASLD, MASH, and advanced liver disease. Participants who lost 15% or more of their body weight were especially protected. These findings suggest that individualized, nutrition-based remote care can help prevent liver disease in people with type 2 diabetes and obesity.

## Introduction

Metabolic dysfunction–associated steatotic liver disease (MASLD) and its more progressive form, metabolic dysfunction–associated steatohepatitis (MASH), are increasingly recognized as major public health concerns in the United States and globally. Recent data indicate that MASLD affects approximately 30–35% of U.S. adults, with prevalence rising to 60–70% among individuals with type 2 diabetes or obesity (1–3). Modeling studies suggest that up to 20% of individuals with MASLD will progress to MASH, corresponding to an estimated 15 million U.S. adults at risk for cirrhosis, liver failure, hepatocellular carcinoma (HCC), and cardiometabolic complications (4). The global economic burden is substantial, with a recent analysis estimating that the clinical, humanistic, and economic impact of MASH exceeds **$**300 billion annually across major countries, underscoring the cost of inaction (5,6). These data are consistent with recent reviews identifying MASLD as a growing global epidemic and one of the leading causes of chronic liver disease worldwide (7,8).

Weight loss of ≥7–10% of body weight is strongly associated with histologic improvements in MASLD/MASH, including resolution of steatohepatitis and regression of fibrosis (9). Accordingly, current guidelines emphasize intentional weight loss as the cornerstone of therapy for MASLD/MASH (10–13). Pharmacologic therapies such as resmetirom (14) and semaglutide (15) are now FDA-recommended options for MASH treatment; however, intentional weight loss remains the primary and most consistently effective strategy for MASH resolution and fibrosis improvement, including when achieved through agents like semaglutide.

The Virta Individualized Nutrition Therapy (VINT), which delivers telemedicine-based remote care, health coaching, and carbohydrate-reduced nutrition therapy with an emphasis on nutritional ketosis, has demonstrated durable weight loss and metabolic improvements across type 2 diabetes, prediabetes, and overweight/obesity populations in both clinical trials and real-world settings (16–20). Post-hoc analyses of the clinical trial data further revealed significant improvements in liver health, including normalization of alanine aminotransaminase (ALT) in 61% of those with elevated baseline values, reductions in aspartate transaminase (AST) and alkaline phosphatase (ALP), and improvements in surrogate MASLD markers. 20% fewer participants met criteria for suspected steatosis and 15% fewer classified as not having suspected fibrosis after one year (21).

A reduction in new-onset liver-related events (LREs) is the endpoint established by the FDA to support full approval for medications such as resmetirom or semaglutide, which are currently only conditionally approved. To address this gap, we analyzed claims data to assess the incidence of new-onset MASLD, the development of new-onset MASH (including cases that may have progressed from a prior MASLD diagnosis), and advanced liver disease among patients enrolled in VINT. For this purpose, we leveraged ICD-10 diagnostic codes to identify clinically meaningful endpoints at the population level.

## Materials and Methods

### Data Source

This study used data from the Komodo Healthcare Map™, a nationwide U.S. commercial administrative claims database capturing pseudonymized medical, pharmacy, and laboratory claims from a large insured population (22). Tokenized demographic records generated via privacy-preserving encryption were linked to enrollment, follow-up, and corresponding claims data for treated participants. A control population of 2.39 million individuals from the same database was used to create the matched control cohort. The August 25, 2025 dataset iteration was analyzed within Sentinel, Komodo Health’s secure real-world evidence platform.

### Study Population, Intervention, and Medication Management

The analysis included adults (≥18 years) with type 2 diabetes, prediabetes, overweight, or obesity who enrolled in the Virta Individualized Nutrition Therapy (VINT) program for diabetes reversal or weight loss between August 2015 and September 2024 and remained enrolled for at least one year. Individuals with contraindications to nutritional ketosis (e.g., type 1 diabetes, active malignancy, pregnancy) were excluded. Full eligibility criteria and intervention details are provided in the Supplementary Methods.

VINT is a fully remote continuous care model combining individualized nutrition therapy, protocol-guided medication management, and health coaching via telemedicine. The dietary approach emphasizes individualized carbohydrate reduction to achieve nutritional ketosis (target β-hydroxybutyrate 0.5–3.0 mmol/L), moderate protein intake (∼1.2–1.5 g/kg ideal body weight/day), and dietary fat to satiety, with a focus on whole, minimally processed foods. Participants receive structured education and ongoing real-time support from licensed medical providers and health coaches.

Medication adjustments were managed by VINT clinicians to ensure safety during carbohydrate reduction, with early dose reductions for hypoglycemia-prone agents (e.g., insulin, sulfonylureas) and individualized titration of other therapies as indicated. Biomarkers, including blood glucose, BHB, and body weight, were monitored daily at initiation, particularly during the first three months, and subsequently tailored to participant preferences and tolerance, with treatment decisions guided by these data and clinician oversight.

### Claims Analysis

For the claims-based analyses, both VINT participants and controls were required to have continuous medical and pharmacy claims coverage defined as no gap greater than 30 days for at least one year prior to the enrollment or index date and at least six months after. Control participants were assigned a randomly generated index date aligned with the distribution of enrollment dates in the VINT cohort to ensure temporal comparability. Baseline measures included in the matching encompassed demographic characteristics (age, sex, and race/ethnicity), clinical comorbidities, medication use, and healthcare costs. Comorbidities were identified using International Classification of Diseases, Tenth Revision, Clinical Modification (ICD-10-CM) codes appearing in either primary or secondary positions within medical claims (see Supplementary Table S1). Several liver conditions were excluded at baseline before matching (details in the supplementary method and Supplementary Table S1). Individuals with MASLD, MASH, and fibrosis were retained to allow evaluation of new-onset and progression outcomes within these at-risk populations. Medication classes assessed at baseline and follow-up are detailed in Supplementary Table S2.

### Study Outcomes

The primary composite outcomes were defined as the time to new onset of any liver related diagnosis, MASH and beyond, advanced liver disease and advanced liver complications (Table 1 for details).

**Table 1.** Definition of Primary Composite Liver Outcomes.

| **Outcome Category** | **Outcome Name** | **Composite Components Included** | **Clinical Rationale** |
| --- | --- | --- | --- |
| **Primary composite outcome** | Any liver-related diagnosis | MASLD; MASH; liver fibrosis; cirrhosis; portal hypertension (PHTN); PHTN complications (ascites, varices, hepatic encephalopathy); hepatocellular carcinoma (HCC); or other abnormal liver imaging findings | Captures the full spectrum of incident liver disease from early to advanced pathology |
| **Primary composite outcome** | MASH and beyond | MASH; liver fibrosis; cirrhosis; PHTN; PHTN complications; HCC | Focuses on progressive liver disease beyond steatosis |
| **Primary outcome of interest** | Advanced liver disease | Cirrhosis; PHTN; PHTN complications; HCC | Represents severe liver disease progression and aligns with clinical trial and regulatory endpoints |
| **Secondary composite outcome** | Advanced liver complications | PHTN; PHTN complications (ascites, varices, hepatic encephalopathy) | Captures clinically significant complications driving morbidity and healthcare utilization |

“New onset” was defined as the first occurrence of the outcome after the index date among participants who met the strategy-specific baseline inclusion criteria. Follow-up was limited to 5 years and was right-censored at the time a liver-related event occurred, at the end of the closed claims period, or at the study end date if no outcome occurred.

Analyses were conducted using three complementary analytic strategies to assess robustness and consistency. Analytic strategy 1 (incident disease cohort) excluded participants with baseline MASLD, MASH, or fibrosis and evaluated all three composite outcomes: any liver-related diagnosis, MASH and beyond, and advanced liver disease. Analytic strategy 2 (incident disease and potential MASLD progression cohort) excluded participants with baseline MASH or fibrosis (allowing baseline MASLD) and evaluated two composite outcomes: MASH and beyond, and advanced liver disease. Analytic strategy 3 (incident disease cohort with maximally restricted baseline) excluded all participants with baseline evidence of liver disease defined as MASLD, MASH, fibrosis, or findings suggestive of liver disease based on imaging, biopsy, or elastography procedure codes and evaluated the any liver-related, MASH-and-beyond and advanced liver disease composite outcomes. For the MASH-and-beyond, advanced liver disease analyses and liver complications, other liver-related diagnoses identified during follow-up (e.g., MASLD, abnormal liver imaging, or inflammatory liver disease) were not counted as events, ensuring a clean assessment of the prespecified composite endpoints.

Secondary outcome analyses were restricted to the VINT cohort. We evaluated whether categorical clinical variables including mean BHB level over 6 months (≥0.3 vs <0.3 mmol/L and ≥0.5 vs <0.5 mmol/L), 6-month weight change (≥10% loss vs <10% loss and ≥15% loss vs <15% loss), and HbA1c change at 1 year (or 6 months if 1-year data were unavailable; ≥0.5 percentage point reduction vs <0.5 point reduction or any increase) were associated with the primary composite outcome of new onset of any liver related diagnosis, as defined in analytic strategy 1.

Exploratory outcomes characterized healthcare utilization and liver-specific procedures (e.g., ultrasound, elastography, CT/MRI, biopsy), comparing intervention participants with matched controls. For the MASLD, MASH, and fibrosis subgroups, we also conducted a descriptive evaluation of concomitant pharmacotherapy among participants with evidence of disease at baseline or who developed MASLD or MASH during follow-up. Medication classes were defined a priori and included metabolic and liver-directed therapies (e.g., GLP-1 receptor agonists, SGLT2 inhibitors, thiazolidinediones [pioglitazone], vitamin E, and statins).

### Statistical analysis

This study was conducted and reported in accordance with the STROBE guidelines for observational research (23), with a completed checklist included in the Supplementary Material. A detailed description of the statistical analysis is in the supplementary methods. Baseline characteristics are summarized as mean (SD) for continuous variables and n (%) for categorical variables. Propensity scores were estimated using the logit of the predicted score, and 1:1 matching without replacement was performed. Exact matching was enforced on sex, age category, race/ethnicity, baseline liver condition indicators (MASLD, MASH, fibrosis), and medications with potential liver benefit, including incretin mimetics and SGLT2 inhibitors. Balance after matching was evaluated using standardized mean differences (SMDs), applying a 0.10 threshold for acceptable balance. Baseline covariates included demographics (age, sex, race/ethnicity), clinical conditions (type 2 diabetes, obesity, hypertension, hyperlipidemia, coronary artery disease, stroke, heart failure, chronic kidney disease stage, albuminuria, smoking, alcohol-related diagnoses, and other cardiovascular disease), liver conditions, baseline liver procedures, medication exposures (SGLT2 inhibitors, GLP-1 receptor agonists, statins, insulin, sulfonylureas, DPP-4 inhibitors, and other liver-beneficial agents such as certain statins, icosapent ethyl, omega-3 acid ethyl esters, vitamin E, α-tocopherol, gemfibrozil, and fenofibrate), one-year pre-index utilization and cost measures, and geographic and socioeconomic indicators (census region and area deprivation index quintile). Between-group balance is described using SMD≤0.10 indicating adequate balance (no hypothesis tests for baseline).

Primary composite time-to-event outcomes (definitions provided in *Study Outcomes*) were analyzed using Cox proportional hazards models to estimate hazard ratios (HRs) and 95% confidence intervals (CIs), comparing VINT participants with matched controls. For matched analyses, stratified Cox models were applied by matched set, with robust standard errors clustered on matched pairs to account for within-set correlation. All models were adjusted for age, sex, and race/ethnicity. Extended models further adjusted for baseline diagnoses of type 2 diabetes and obesity, as well as follow-up use of cardiometabolic medications with potential hepatic effects, including SGLT2 inhibitors, GLP-1 receptor agonists, statins, and other relevant agents. Medication use was modeled both as categorical ever/never exposure and as continuous proportion of days covered (PDC) to capture adherence.

Kaplan–Meier cumulative incidence curves were generated to visualize differences between groups, with numbers at risk displayed at prespecified time points. Incidence rates were calculated as events per 1,000 person-years. Absolute risk reduction (ARR) was computed as the difference in incidence rates between the UC and VINT groups, and the number needed to treat (NNT) was derived as the reciprocal of ARR (1/ARR), representing the number of individuals who would need to receive VINT for one year to prevent one additional new-onset liver diagnosis compared with UC.

To assess the robustness of the findings, several sensitivity analyses were conducted (details in the supplementary method), advanced liver disease outcomes reassessed after a) excluding participants who developed other liver diseases during follow-up, b) survival model adjusted for healthcare utilization variables, c) a Fine Gray sub distribution hazards model, including death from any cause as a competing risk. Additionally, to further evaluate the consistency of all the primary results, analyses were performed using inverse probability of treatment weighting (IPTW) with stabilized weights. Weighted Cox proportional hazards models with robust variance estimators were then used to estimate hazard ratios (HRs) in the pseudo-population generated by IPTW.

Within the VINT cohort, secondary outcomes were assessed using multiple Cox models examining associations of weight loss, ketone exposure (BHB), and HbA1c change with incident liver disease, with additional Poisson and trend analyses across weight-loss categories. Exploratory analyses descriptively evaluated liver disease state transitions, liver-specific imaging/procedures, and medication use patterns in relevant subcohorts; full methodological details are provided in the Supplementary Methods.

All statistical tests were two-sided (α = 0.05) with 95% confidence intervals (CIs), and analyses were conducted using R software (R packages used are listed in the Supplementary statistical method section).

## Results

### Study Participants Flow

Within the Komodo Sentinel claims environment, the initial study population included 78,755 individuals in the intervention cohort and 2.39 million in the control cohort (Figure 1). After applying the closed claims requirement, 10,170 intervention participants and 509,727 controls remained. Following the additional eligibility criteria of at least one year of continuous program enrollment, a minimum of six months of claims data follow up and after excluding some baseline comorbidities or conditions, 5,236 intervention participants and 200,778 controls met inclusion requirements and were retained for propensity score matching (Figure 1; Supplementary Table S3).

**Figure 1.**
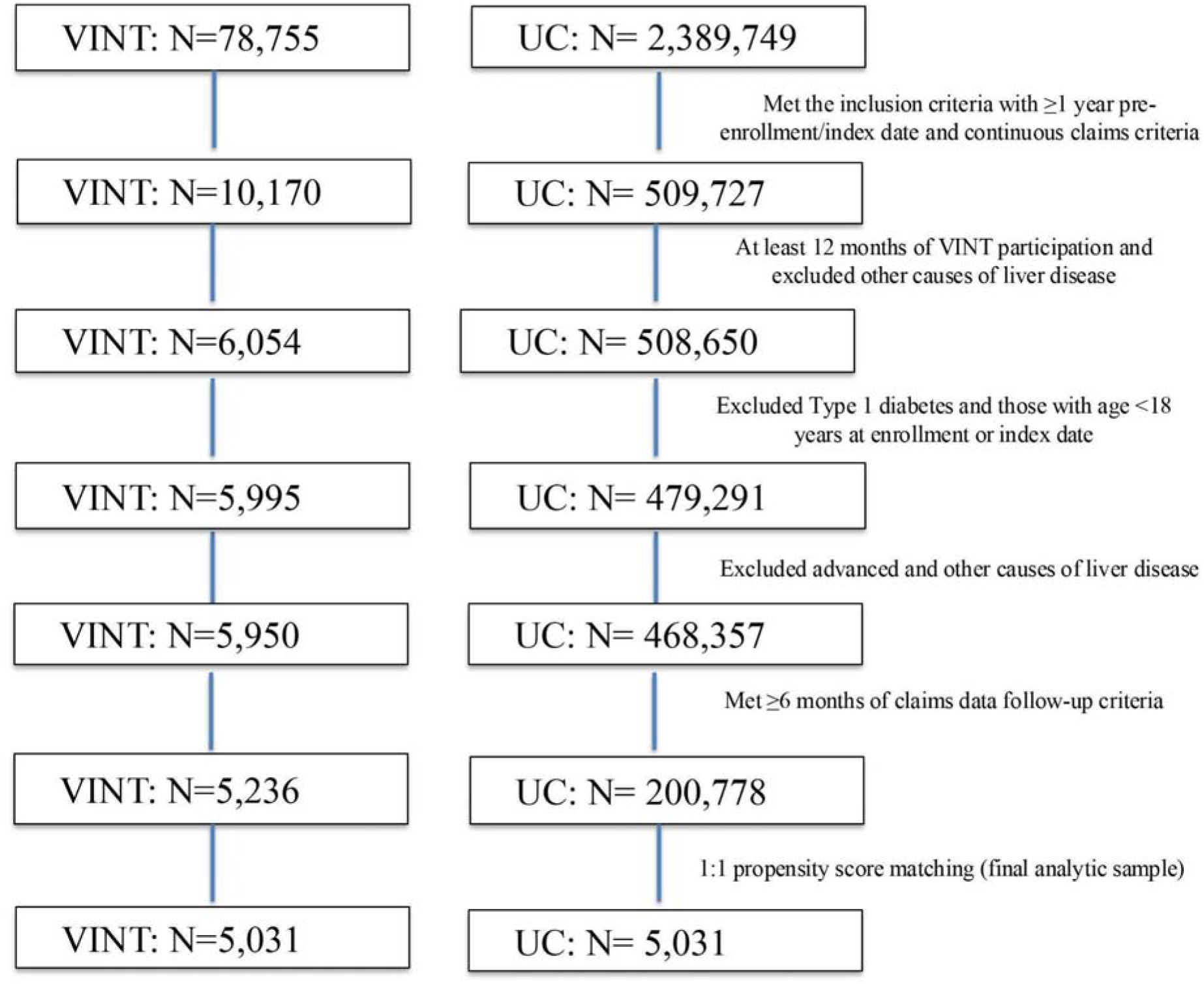
Flow diagram depicting patient selection and attrition from the initial Komodo Healthcare Sentinel cohort to the final matched analytic sample.

### Baseline Characteristics

After 1:1 propensity score matching, the analytic sample consisted of 5,031 VINT participants and 5,031 controls (usual care, UC) (Figure 1; Tables 2 and 3), representing roughly 10% of the original dataset with similar tenure. To assess representativeness, the matched VINT cohort was compared with VINT participants excluded during screening. Baseline demographics and aggregated variables were broadly similar between groups, with most standardized mean differences (SMDs) ≤ 0.1. Minor imbalances were noted for race and ethnicity (SMD = 0.29) and program type (diabetes reversal vs. sustainable weight loss, SMD = 0.18) (Supplementary Table S4). Although these values exceed the stringent 0.1 threshold used in this study, they remain close to the more permissive 0.2 threshold that is sometimes applied in observational research to indicate acceptable balance. Post-matching, baseline characteristics were well balanced between VINT and UC across all covariates used in the matching (all SMDs ≤0.1) as well as most comorbidities, clinical conditions, and medications not directly included in the matching process, except for follow-up duration (Tables 2 and 3). The matched cohorts had a mean age of 53 years, and the distribution of males and females was similar between the groups. At baseline, 5.1% of participants had MASLD/MASH (including fibrosis), 51% had type 2 diabetes, and 36% had obesity. Mean follow-up was 2.2 years in the VINT group and 1.9 years in the UC group.

**Table 2.** Baseline demographics, disease trajectory as assessed by cost, and area level variables of propensity score 1:1 matched cohort of VINT and UC.

| Characteristics | VINT (N=5,031) | UC (N=5,031) | SMD |
| --- | --- | --- | --- |
|  | Mean (SD) or N (%) | Mean (SD) or N (%) |  |
| Demographics |  |  |  |
| Age at registration | 52.9 (9.3) | 53.0 (9.5) | 0.01 |
| Follow-up duration | 804.8 (415.9) | 684.8 (492.4) | 0.26 |
| Gender |  |  | <0.001 |
| Female | 2853 (56.7) | 2853 (56.7) |  |
| Male | 2161 (43.0) | 2161 (43.0) |  |
| Unknown | 17 (0.3) | 17 (0.3) |  |
| Race/Ethnicity |  |  | <0.001 |
| White | 2682 (53.3) | 2682 (53.3) |  |
| Black or African American | 533 (10.6) | 533 (10.6) |  |
| Hispanic or Latino | 382 (7.6) | 382 (7.6) |  |
| Asian or Pacific Islander | 162 (3.2) | 162 (3.2) |  |
| Other | 98 (1.9) | 98 (1.9) |  |
| Unknown | 1174 (23.3) | 1174 (23.3) |  |
| Baseline Costs |  |  |  |
| Total Cost of Care | 13335.2 (25726.3) | 14970.6 (31641.8) | 0.06 |
| Outpatient Cost | 8020.8 (18317.3) | 8799.8 (25037.2) | 0.04 |
| Inpatient Cost | 1285.3 (9523.0) | 1362.2 (7272.6) | 0.01 |
| Pharmacy Cost | 4013.3 (11428.0) | 4785.3 (13714.8) | 0.06 |
| Baseline area level variables |  |  |  |
| Region |  |  | 0.10 |
| Midwest | 1964 (39.0) | 1721 (34.2) | 0.10 |
| Northeast | 482 (9.6) | 603 (12.0) | 0.08 |
| South | 2055 (40.8) | 2043 (40.6) | 0.01 |
| West | 530 (10.5) | 664 (13.2) | 0.09 |
| Area Deprivation Index (ADI) Quintiles |  |  | 0.08 |
| ADI 1 | 825 (16.4) | 982 (19.5) | 0.08 |
| ADI 2 | 2084 (41.4) | 1924 (38.2) | 0.06 |
| ADI 3 | 1401 (27.8) | 1322 (26.3) | 0.04 |
| ADI 4 | 463 (9.2) | 516 (10.3) | 0.04 |
| ADI 5 | 258 (5.1) | 287 (5.7) | 0.03 |

**Note:** Standardized mean differences (SMDs) were calculated using the TableOne package in R. For continuous variables, *p*-values were derived using the student's *t*-test when the normality assumption was met and the Wilcoxon rank-sum test otherwise. For categorical variables, *p*-values were obtained using the chi-square test or Fisher's exact test when cell counts were small. SMDs were computed as the absolute difference in means (or proportions for categorical variables) between groups divided by the pooled standard deviation. SMDs were calculated using both the TableOne and MatchIt packages to cross-validate covariate balance estimates. To evaluate covariate balance between matched groups, $SMD \leq 0.1$ was used as the criterion for acceptable balance. Unlike *p*-values, which are influenced by sample size and may indicate statistically significant differences that are not clinically meaningful in large samples, SMD provides a sample size-independent measure of balance that quantifies the magnitude of difference between groups. Therefore, SMDs were used as the primary metric for assessing post-matching balance, while *p*-values were reported descriptively.
**Abbreviations:** UC, usual care; VINT, Individualized Nutrition Therapy (intervention) program

**Table 3.** Baseline comorbidities and medication use of propensity score 1:1 matched cohort of VINT and UC.

| Characteristics | VINT (N=5,031) | UC (N=5,031) | SMD |
| --- | --- | --- | --- |
|  | Mean (SD) or N (%) | Mean (SD) or N (%) |  |
| Baseline comorbidities |  |  |  |
| Type 2 Diabetes | 2552 (50.7) | 2552 (50.7) | <0.001 |
| Obesity | 1831 (36.4) | 1831 (36.4) | <0.001 |
| MASLD or MASH | 255 (5.1) | 255 (5.1) | <0.001 |
| MASH | 2 (0.0) | 2 (0.0) | <0.001 |
| MASLD | 254 (5.0) | 254 (5.0) | <0.001 |
| Liver procedure | 170 (3.4) | 170 (3.4) | <0.001 |
| Fibrosis | 1 (0.0) | 1 (0.0) | <0.001 |
| Coronary Artery Disease | 296 (5.9) | 253 (5.0) | 0.04 |
| Any other cardiovascular disease | 41 (0.8) | 47 (0.9) | 0.01 |
| Stroke | 80 (1.6) | 85 (1.7) | 0.01 |
| Atrial Fibrillation | 97 (1.9) | 137 (2.7) | 0.05 |
| Heart Failure | 63 (1.3) | 80 (1.6) | 0.03 |
| Hyperlipidemia | 2616 (52.0) | 2595 (51.6) | 0.01 |
| Hypertension | 2562 (50.9) | 2570 (51.1) | 0.003 |
| Chronic Kidney Disease (CKD) | 279 (5.5) | 301 (6.0) | 0.02 |
| CKD Stage 3 and beyond | 76 (1.5) | 58 (1.2) | 0.05 |
| Albuminuria | 121 (2.4) | 118 (2.3) | 0.004 |
| Cancer | 260 (5.2) | 280 (5.6) | 0.02 |
| Peripheral Arterial Disease | 101 (2.0) | 163 (3.2) | 0.08 |
| Sleep Disorder | 1375 (27.3) | 1261 (25.1) | 0.05 |
| Smoking | 162 (3.2) | 230 (4.6) | 0.07 |
| Alcohol use/abuse diagnosis | 46 (0.9) | 64 (1.3) | 0.03 |
| Baseline medications |  |  |  |
| Potential liver benefit medications or supplements | 2032 (40.4) | 2032 (40.4) | <0.001 |
| Omega-3 fatty acids | 37 (0.7) | 43 (0.9) | 0.01 |
| Fibrates | 159 (3.2) | 173 (3.4) | 0.02 |
| Vitamin E | 1 (0.0) | 2 (0.0) | 0.01 |
| Obetocholic Acid | 0 (0.0) | 0 (0.0) | NA |
| Incretin Mimetics | 975 (19.4) | 975 (19.4) | <0.001 |
| Semaglutide | 465 (9.2) | 527 (10.5) | 0.04 |
| Tirzepatide | 86 (1.7) | 164 (3.3) | 0.100 |
| Liraglutide | 118 (2.3) | 110 (2.2) | 0.01 |
| Dulaglutide | 393 (7.8) | 300 (6.0) | 0.07 |
| Exenatide | 57 (1.1) | 48 (1.0) | 0.02 |
| Lixisenatide | 23 (0.5) | 12 (0.2) | 0.04 |
| Albiglutide | 1 (0.0) | 3 (0.1) | 0.02 |
| SGLT2i | 572 (11.4) | 572 (11.4) | <0.001 |
| Insulin | 447 (8.9) | 494 (9.8) | 0.03 |
| DPP4i | 292 (5.8) | 292 (5.8) | <0.001 |
| Sulfonylurea | 267 (5.3) | 241 (4.8) | 0.02 |
| TZD | 142 (2.8) | 134 (2.7) | 0.01 |
| Other T2D medications | 2167 (43.1) | 2104 (41.8) | 0.03 |
| Anticoagulant | 133 (2.6) | 252 (5.0) | 0.12 |
| Antiplatelet | 182 (3.6) | 250 (5.0) | 0.07 |
| Beta Blocker | 763 (15.2) | 921 (18.3) | 0.08 |
| Calcium Channel Blocker | 712 (14.2) | 826 (16.4) | 0.06 |
| Diuretic | 1206 (24.0) | 1229 (24.4) | 0.01 |
| Other Antihypertensive | 123 (2.4) | 218 (4.3) | 0.10 |
| Statin | 2218 (44.1) | 2218 (44.1) | <0.001 |
| Other Lipid Lowering | 313 (6.2) | 323 (6.4) | 0.01 |
| PCSK9i | 18 (0.4) | 9 (0.2) | 0.04 |

**Note:** Standardized mean differences (SMDs) were calculated using the TableOne package in R. For continuous variables, *p*-values were derived using the student's *t*-test when the normality assumption was met and the Wilcoxon rank-sum test otherwise. For categorical variables, *p*-values were obtained using the chi-square test or Fisher's exact test when cell counts were small. SMDs were computed as the absolute difference in means (or proportions for categorical variables) between groups divided by the pooled standard deviation. SMDs were calculated using both the TableOne and MatchIt packages to cross-validate covariate balance estimates. To evaluate covariate balance between matched groups, $SMD \leq 0.1$ was used as the criterion for acceptable balance. Unlike *p*-values, which are influenced by sample size and may indicate statistically significant differences that are not clinically meaningful in large samples, SMD provides a sample size-independent measure of balance that quantifies the magnitude of difference between groups. Therefore, SMDs were used as the primary metric for assessing post-matching balance, while *p*-values were reported descriptively.
**Abbreviations:** UC, usual care; VINT, Individualized Nutrition Therapy (intervention) program; T2D, type 2 diabetes; MASLD, metabolic dysfunction-associated steatotic liver disease; MASH, metabolic dysfunction-associated steatohepatitis; CKD, chronic kidney disease; SGLT2i, sodium-glucose cotransporter-2 inhibitor; DPP4i, dipeptidyl peptidase-4 inhibitor; RAASi, renin-angiotensin-aldosterone system inhibitor (e.g., ACEi/ARB/ARNI); MRA, mineralocorticoid receptor antagonist (steroidal); non-MRA, non-steroidal mineralocorticoid receptor antagonist (e.g., finerenone); PCSK9i, proprotein convertase subtilisin/kexin type 9 inhibitor, potential drugs with liver benefits included Vitamin E, omega 3 fatty acids, fibrates, atorvastatin, rosuvastatin, and pravastatin

Although the mean follow-up duration was shorter in the UC group, survival analyses inherently accommodate unequal follow-up by using time-to-event with right censoring; thus, estimates remain unbiased under non-informative censoring. Furthermore, the selection of index dates for UC was aligned with the distribution of VINT enrollment dates. This approach ensured that seasonal patterns, temporal shifts, or year-specific effects were accounted for, as illustrated by the overlapping distribution of index dates between groups (Supplementary Figure 1).

### Primary outcomes

Across all analytic strategies, VINT participants had consistently lower incidence rates of new-onset any liver-related diagnoses compared with UC. Under strategy 1, the rates were 27.3 vs 42.8 per 1,000 person-years (HR = 0.61; 95% CI 0.52–0.72; *p* < 0.001; Figure 2A), and under strategy 3, 27.4 vs 42.4 (HR = 0.62; 95% CI 0.52–0.73; *p* < 0.001; Figure 2B). These associations remained significant and of similar or greater magnitude after adjustment for medication use (Table 4). Based on incidence rates, the corresponding absolute risk reductions were approximately 15 fewer events per 1,000 person-years, yielding NNTs of 65–67 per year.

**Figure 2.**
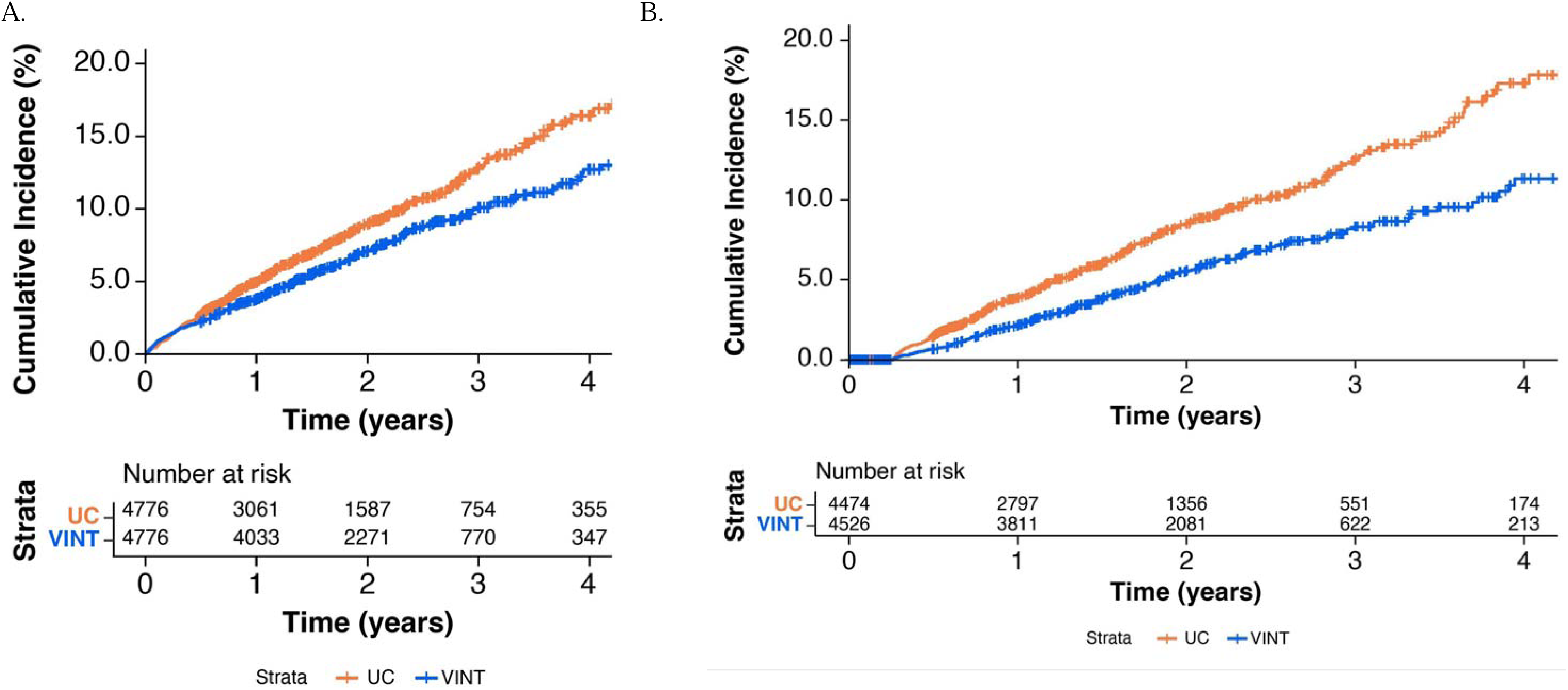
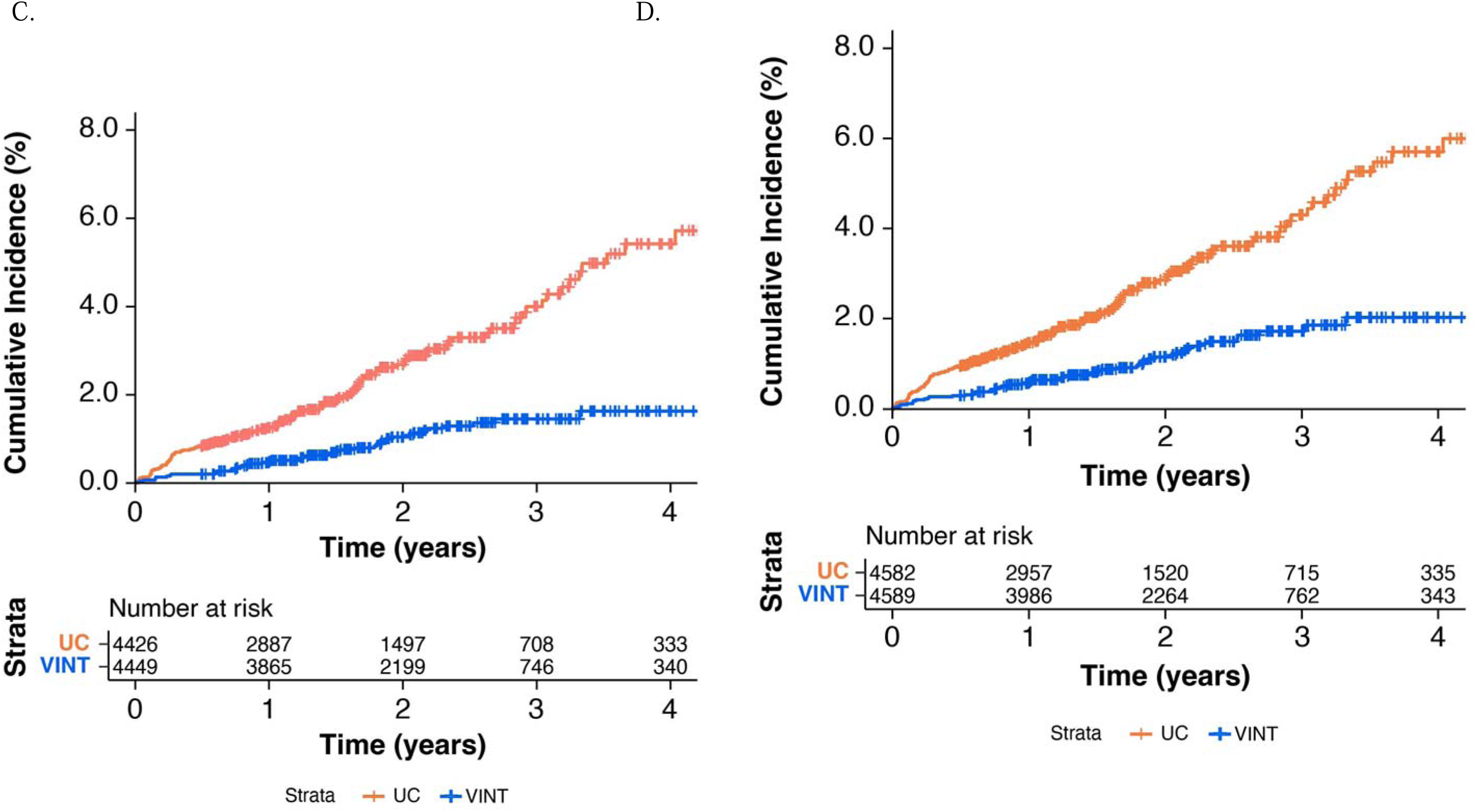
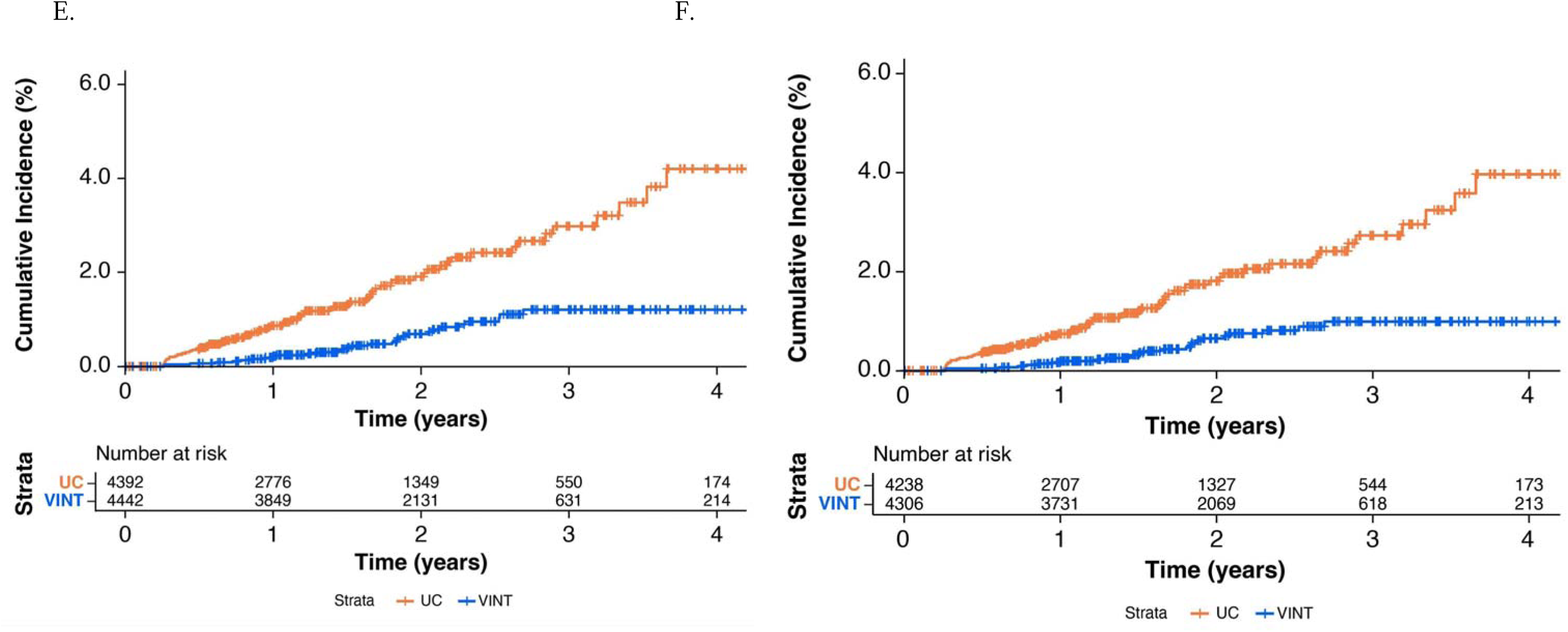
Cumulative incidence curves for new-onset of all liver diagnoses under (A) analytic strategy 1 (incident disease cohort), (B) analytic strategy 3 (incident disease cohort with maximally restricted baseline), new-onset of MASH and beyond under (C) analytic strategy 1 (incident disease cohort), (D) analytic strategy 2 (incident disease cohort and potential baseline MASLD progression), new-onset of fibrosis, cirrhosis and/or portal hypertension (E) analytic strategy 1 (incident disease cohort), (F) analytic strategy 2 (incident disease cohort and potential baseline MASLD progression). Curves are displayed up to 4 years to aid interpretability, given the decreasing sample size late in follow-up, though all analyses were conducted using the full observation window with standard right-censoring.

**Table 4.**
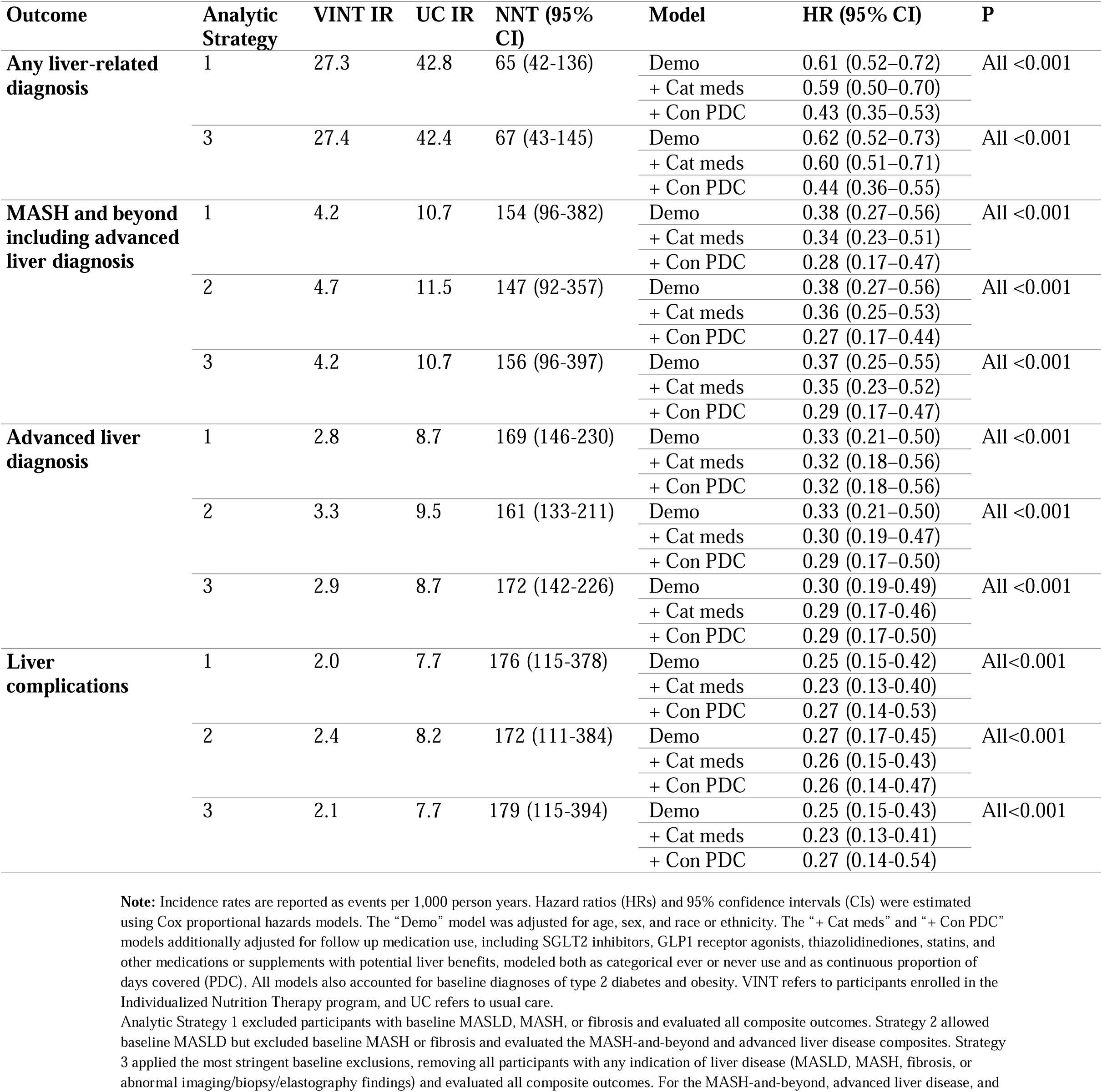
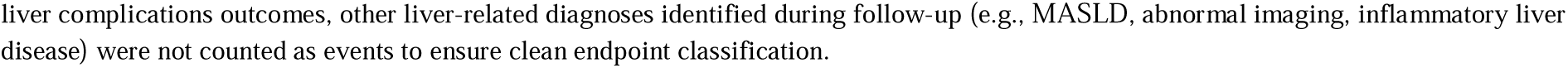
Incidence rates, number needed to treat and hazard ratios for new-onset liver diagnoses, including MASH, advanced disease, fibrosis, cirrhosis and portal hypertension, under different analytic strategies.

For new-onset MASH and beyond, incidence rates were consistently lower among VINT participants across all three analytic strategies: 4.2 vs 10.7 per 1,000 person-years (HR = 0.38; 95% CI 0.27–0.56; *p* < 0.001; Figure 2C) under strategy 1; 4.7 vs 11.5 (HR = 0.38; 95% CI 0.27–0.56; *p* < 0.001; Figure 2D) under strategy 2; and 4.2 vs 10.7 (HR = 0.37; 95% CI 0.25–0.55; *p* < 0.001) under strategy 3. These findings remained significant and directionally consistent following adjustment for medication use (Table 4). Based on incidence rates, absolute risk reductions ranged from 6.4–6.8 events per 1,000 person-years, corresponding to NNTs of 147–156 per year.

For new-onset advanced liver disease, VINT participants again demonstrated substantially lower incidence rates: 2.8 vs 8.7 per 1,000 person years (HR = 0.33; 95% CI 0.21–0.50; *p* < 0.001; Figure 2E) under strategy 1; 3.3 vs 9.5 (HR = 0.33; 95% CI 0.21–0.50; *p* < 0.001; Figure 2F) under strategy 2; and 2.9 vs 8.7 (HR = 0.30; 95% CI 0.19–0.49; *p* < 0.001) under strategy 3. The corresponding absolute risk reductions were 5.8-6.2 fewer events per 1,000 person years, yielding NNTs of 169-172 per year. As summarized in Table 4, the hazard ratios remained consistent after adjustment for medication exposure. Sensitivity analyses, including the exclusion of participants who developed other causes of liver disease during follow up and a Fine Gray subdistribution hazards model with death as a competing event, yielded similar results, further supporting the robustness of the findings for advanced liver disease (Supplementary Results). Another set of sensitivity analyses adjusting for healthcare utilization measures including outpatient costs, imaging frequency, laboratory testing, and evaluations per member per year produced results consistent with the primary analysis, reinforcing that the observed differences in advanced liver disease between VINT and UC were not attributable to differences in diagnostic activity or healthcare contact.

For new-onset liver complications, VINT participants again demonstrated substantially lower incidence rates compared with controls: 2.0 vs 7.7 per 1,000 person years (HR = 0.25; 95% CI 0.15–0.42; *p* < 0.001) under strategy 1; 2.4 vs 8.2 (HR = 0.27; 95% CI 0.17–0.45; *p* < 0.001) under strategy 2; and 2.1 vs 7.7 (HR = 0.25; 95% CI 0.15–0.43; *p* < 0.001) under strategy 3. The corresponding absolute risk reductions were approximately 5.6 to 5.8 fewer events per 1,000 person years, yielding numbers needed to treat (NNTs) ranging from 172 to 179 per year.

Diagnostic distribution at follow-up events (Figures 3A–3C; overall cohort by analytic strategy) showed UC had a greater proportion of advanced/later-stage liver diagnoses than VINT. In a separate subcohort with baseline MASLD/MASH/fibrosis and a follow-up date-coded diagnosis (Supplementary Table S5), most participants remained in the same diagnostic category, with higher stability in VINT (74.3%) than UC (69.6%); transitions to nonspecific or advanced diagnoses were infrequent and were similar or slightly lower in VINT.

**Figure 3.**
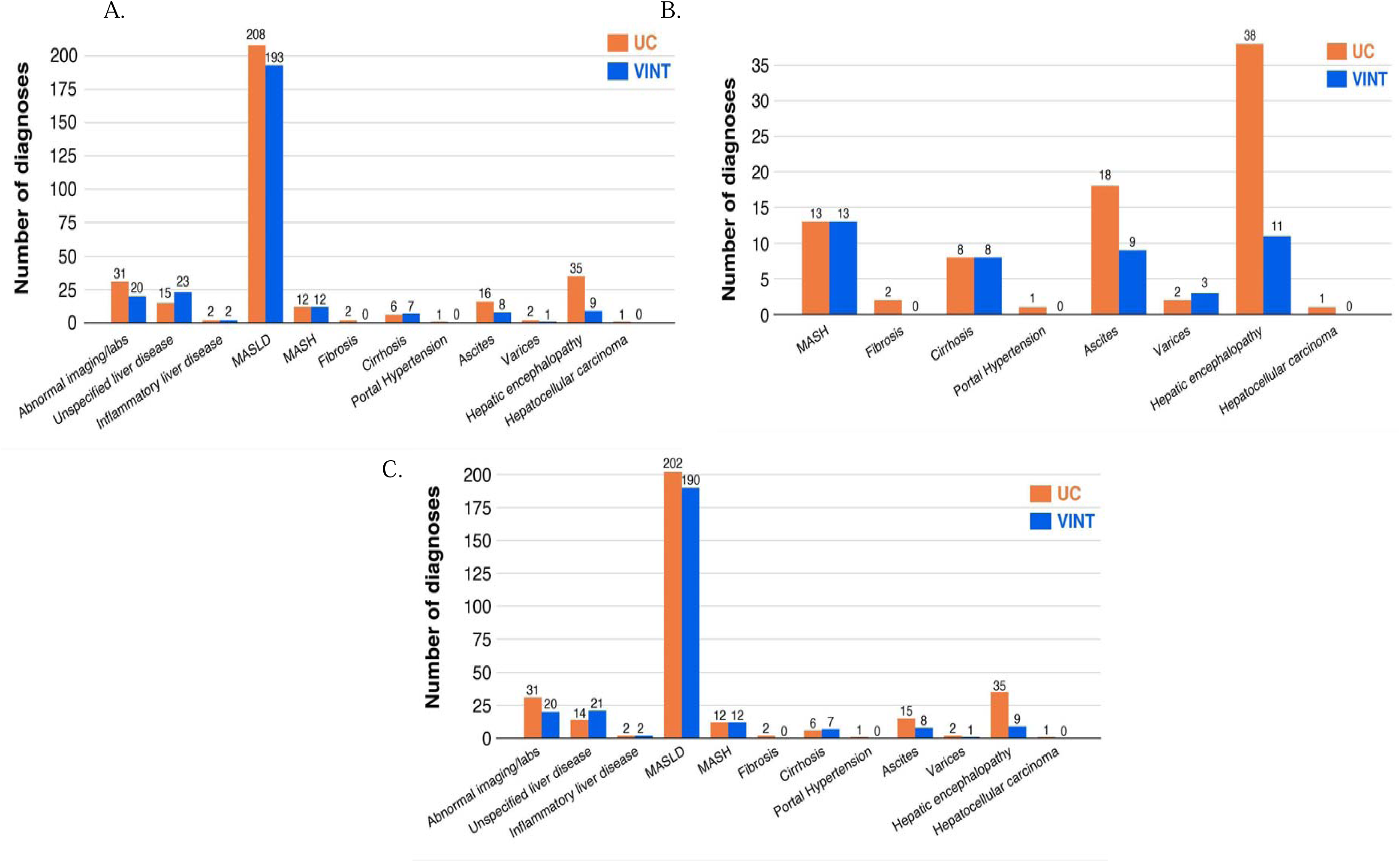
Distribution of liver diagnoses observed after index or enrollment, compiled as events contributing to the primary composite outcomes under (A) Analytic Strategy 1, (B) Analytic Strategy 2, and (C) Analytic Strategy 3.

Sensitivity analyses using IPTW-weighted Cox regression produced consistent results, which are presented in Supplementary Table S6.

### Secondary and exploratory outcomes (see detailed results in the Supplementary Results)

In the VINT cohort, ≥15% weight loss was associated with a significantly lower risk of incident liver diseases, with a clear dose-response across weight-loss categories, associations were attenuated after medication adjustment (Figures 4A and 4B). Mean BHB and HbA1c reduction were not associated with incident liver disease. Exploratory analyses showed fewer liver-specific imaging/procedures in VINT versus usual care despite similar overall healthcare utilization, and medication use patterns were comparable between groups (Supplementary Tables S7 and S8).

**Figure 4.**
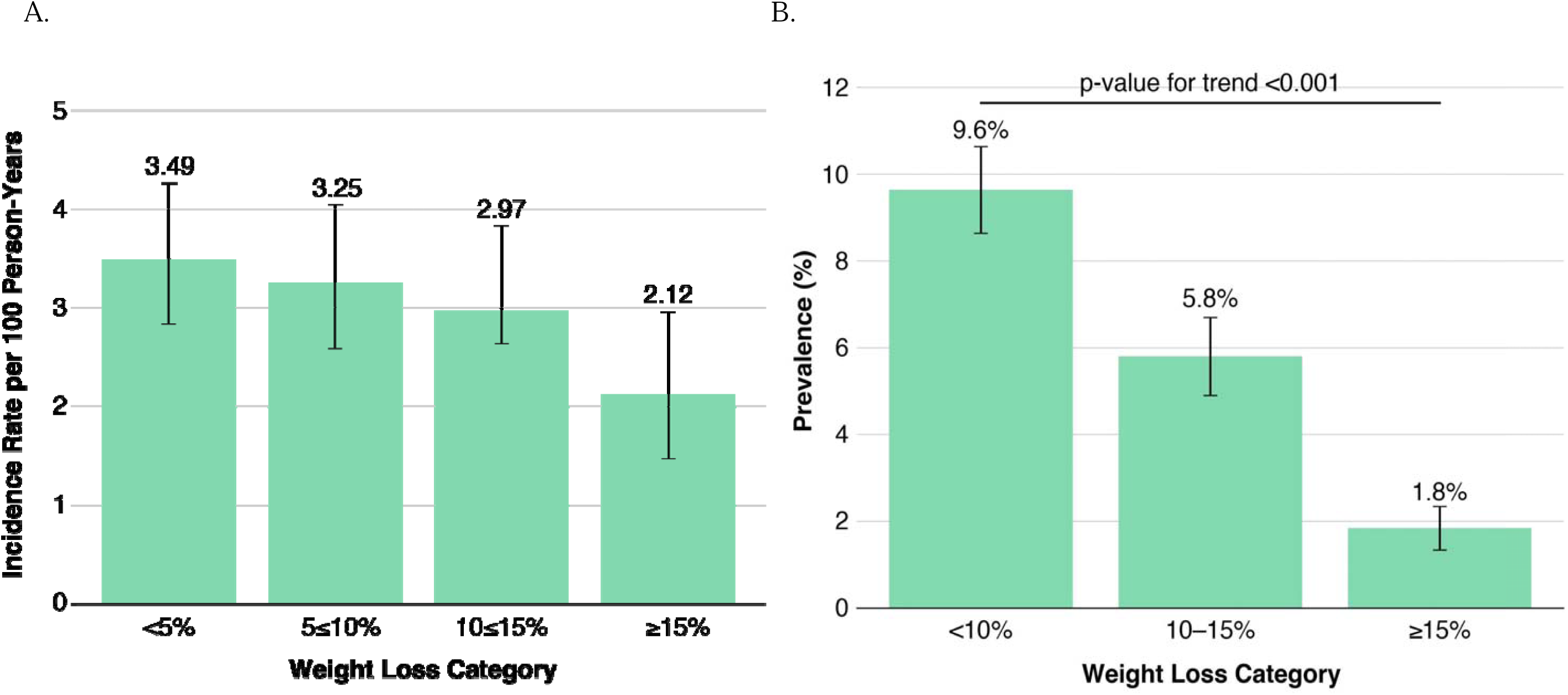
Dose-response relationship between the level of weight loss category in VINT versus UC in (A) incidence rates of any liver diagnoses onset (B) prevalence of any liver diagnoses during follow-up

## Discussion

In this large US based matched cohort of adults at risk for MASLD, participants enrolled in the VINT program was associated with a lower risk of liver related events (Figures 5A and 5B), over five years. The NNT, approximately 170 to prevent one case of advanced liver disease and 180 to prevent one liver complication per year, highlights the meaningful population level impact achievable with individualized nutrition focused care. The VINT model, already deployed at scale for type 2 diabetes and obesity care in the United States, has immediate relevance for liver disease prevention. These findings support earlier MASLD identification and routine liver assessment in high-risk populations (24–26), integration of nutrition-aligned metabolic therapies with pharmacologic treatment, and policy efforts addressing upstream metabolic drivers such as food insecurity and equitable access to nutrition-centered care (27, 28).

**Figure 5.**
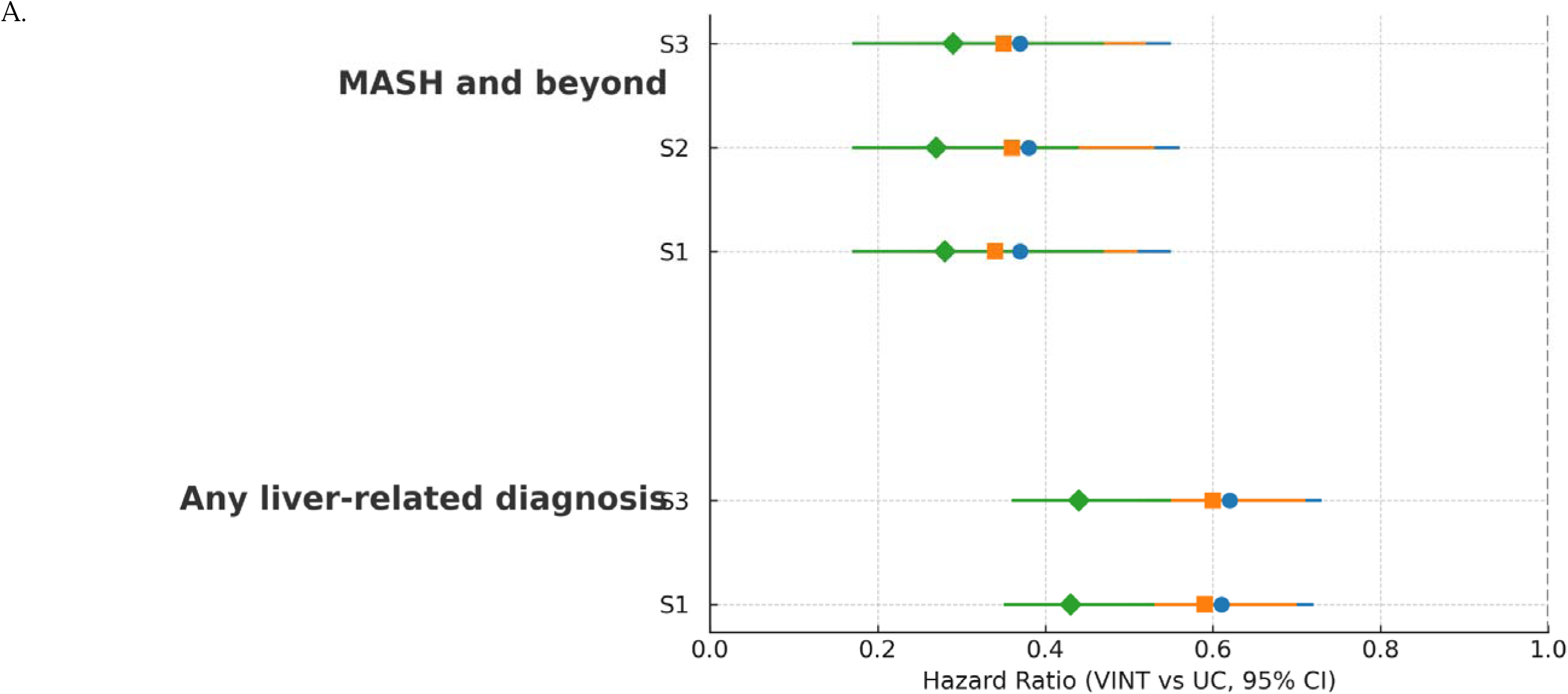
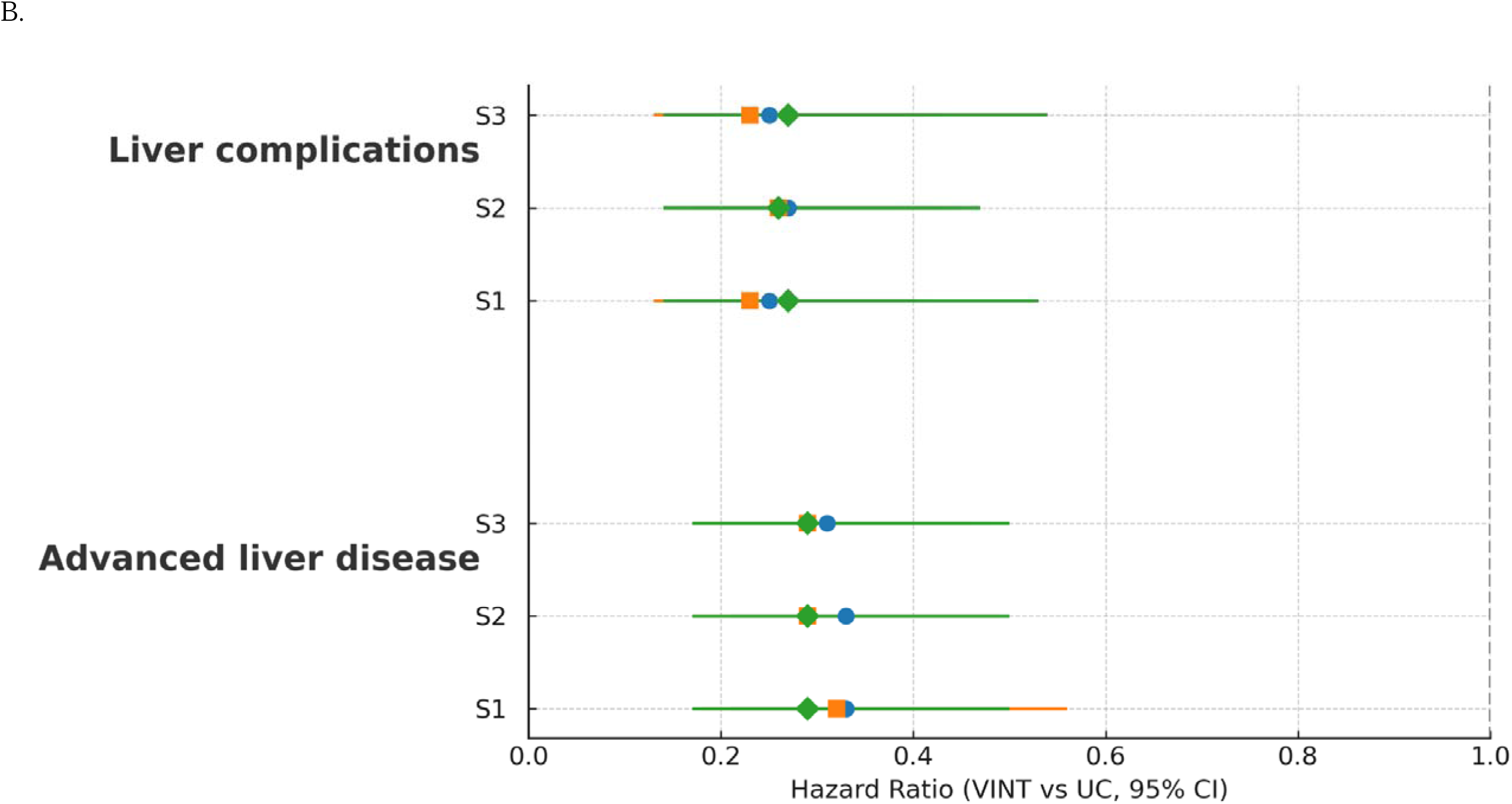
Forest plot displaying hazard ratios (HRs) and 95% confidence intervals (CIs) for the association between participation in the VINT program and the incidence of (A) any liver-related diagnosis (S1 and S3) and MASH and beyond under three analytic strategies (S1–S3) (B) advanced liver disease and liver complications under three analytic strategies (S1–S3) Estimates are shown for three model specifications: the demographic-adjusted model (“Demo”), the model additionally adjusted for follow-up medication use (“Cat meds”), and the model adjusted for continuous proportion of days covered (PDC) with medications of potential liver benefit (“Con PDC”). Vertical dashed line at HR = 1.0 denotes no association. HRs < 1 indicate lower risk among VINT participants compared with usual care.

Pharmacologic studies have also examined primary prevention of liver disease in diabetes populations. In a Veterans Health Administration cohort, GLP 1 receptor agonists were associated with lower risks of cirrhosis, its complications, and liver related mortality compared with DPP 4 inhibitors (29), and a Korean population study found that thiazolidinediones reduced risk of a composite MASLD or MASH outcome compared with DPP 4 inhibitors (30). While these findings highlight the role of medications in liver disease prevention, our results extend this evidence to a scalable, non-pharmacologic, lifestyle-based model. Although advanced liver disease events were infrequent, likely reflecting earlier disease stage, limited follow up, and the slow progression of MASLD and MASH (31, 32), VINT participation was associated with statistically significant reductions in new onset liver related events, including fibrosis, cirrhosis, portal hypertension, and related complications. These patterns parallel those seen in GLP 1 receptor agonist studies and emphasize the potential of early lifestyle interventions to alter liver disease trajectories in real world primary prevention settings. Importantly, medication use was balanced between cohorts, indicating that the observed benefits are independent of pharmacologic therapy.

Weight loss is a well-established therapy for improving MASLD, MASH, and fibrosis and remains central to all major guideline recommendations (9–13). VINT’s continuous remote care model has demonstrated clinically meaningful and durable weight loss among adults with type 2 diabetes and overweight or obesity, with participants achieving and maintaining approximately 10–13 percent loss at two years in clinical trials (16–18) and 9–10 percent in real world settings (19, 20). Consistent with the landmark findings of Vilar Gómez et al., where ten percent or greater weight loss produced the highest rates of MASH resolution and fibrosis regression (9), our analysis showed that fifteen percent or greater weight loss was significantly associated with a lower risk of new onset liver disease. We also observed a clear dose response relationship between the degree of weight loss and both incidence and prevalence of liver disease. These findings align with a recent Cleveland Clinic cohort study of 110 patients showing that greater than five percent weight loss reduced the risk of progression to cirrhosis among individuals with obesity and MASH related F3 fibrosis (33). Collectively, these results highlight that both the magnitude and durability of weight loss are key to preventing liver disease progression at the population level.

Sustaining weight loss remains challenging due to limited time, resources, and nutrition support in routine care (34–38). In contrast, VINT has demonstrated long term durability, maintaining approximately 8 percent weight loss over five years in adults with type 2 diabetes (39, 40) and 7–13 percent over four years in real world cohorts (20). Sustained weight loss is critical, as improvements in steatosis, MASH resolution, and fibrosis regression are often lost with weight regain. Importantly, VINT supports individuals using incretin-based therapies such as semaglutide, including deprescription pathways that help maintain long term weight loss, an important feature given the high real world discontinuation rate of GLP 1 therapies (41). Consistent with these findings, the Cleveland Clinic study similarly emphasized that weight loss greater than 5 percent must be sustained for at least one year to meaningfully reduce progression from MASH related F3 fibrosis to cirrhosis (33). Although ketone levels were not independently associated with reduced risk of new onset liver disease in this analysis, prior evidence indicates that nutritional ketosis supports long term adherence, weight loss durability, and may potentially contribute to MASLD and MASH improvement through multiple mechanisms including inhibition of the NLRP3 inflammasome, improved insulin sensitivity, reduced hepatic de novo lipogenesis, and epigenetic modulation (42–50). Longer term assessments of sustained ketosis are warranted.

Recently approved pharmacotherapies such as semaglutide and resmetirom represent important advances in MASLD and MASH management. Semaglutide produces robust weight loss and MASH improvement, while resmetirom provides direct hepatic benefits including histologic improvement in inflammation and fibrosis (14, 15, 51). Responses to resmetirom appear more pronounced when clinically meaningful weight loss is achieved (52), underscoring the additive potential of combining pharmacotherapy with structured lifestyle-based care. While these agents may ultimately be used together (53), embedding them within a lifestyle first framework could maximize clinical benefit (54) and potentially reduce long term reliance on dual therapy. Importantly, regulatory agencies require post approval evaluation of pharmacotherapies on liver related events such as progression to cirrhosis, development of portal hypertension, hepatocellular carcinoma, liver related hospitalization, and liver specific mortality (55).

This study has several strengths, including its large real world matched cohort design with long term follow up and the use of multiple complementary analytic approaches to strengthen causal inference. The robustness of the findings was supported by consistent results across propensity score matching, inverse probability of treatment weighting, and three analytic strategies. Multiple sensitivity analyses further supported the reliability of the results. Excluding participants who developed other causes of liver disease during follow up yielded similar estimates for the advanced liver disease outcome. Adjusting for healthcare utilization measures produced consistent results, indicating that differences in liver outcomes between VINT and UC were not driven by variation in healthcare contact or diagnostic activity. To account for potential competing risk from mortality, a Fine Gray subdistribution hazards model confirmed that the cumulative incidence of advanced liver disease remained significantly lower among VINT participants even when death was included as a competing event.

Several limitations should be considered, with additional details provided in the Supplementary Discussion. MASLD identification at baseline and follow-up relied on ICD-10 codes due to the absence of anthropometric, laboratory, and imaging data in the claims-based Komodo database, precluding inclusion of measures such as weight, BMI, A1c, and lipid profiles. This may have led to under-ascertainment or misclassification of MASLD and MASH. The requirement for one year of continuous enrollment ensured data completeness but likely selected for individuals with stable insurance and greater healthcare engagement, as only 10–15% of claims populations meet such criteria. VINT participants completing at least one year may also represent a more adherent subgroup, though this duration ensured sufficient exposure and follow-up. Lower rates of liver imaging among VINT participants likely reflect improved metabolic health rather than reduced surveillance, supported by sensitivity analyses showing comparable overall healthcare utilization, imaging, medication use, and evaluation frequency between groups. Mortality analyses were limited to all-cause deaths captured within the claims window, with no cause-of-death data available. The relatively short mean follow-up may have limited detection of advanced hepatic outcomes. Ketone exposure was assessed only in the first six months, potentially underrepresenting long-term nutritional ketosis. Finally, the predominantly commercially insured population and lack of income data may limit generalizability to lower-income or more diverse groups, although neighborhood deprivation was similar between cohorts and residual confounding cannot be ruled out.

Taken together, these findings demonstrate that participation in a structured, individualized nutrition program emphasizing carbohydrate reduction, nutritional ketosis, and continuous remote care was associated with a lower risk of incident MASLD, MASH, and advanced liver diseases including fibrosis, cirrhosis, portal hypertension, and related complications. To our knowledge, this is the first study to demonstrate that FDA recommended liver endpoints decrease and are achievable by non-pharmacological, non-surgical interventions. Collectively, the results support the potential for durable, lifestyle first models to alter the clinical trajectory of liver disease and reduce system level burden and highlight the need for future studies evaluating integration with emerging therapies, and long term cost effectiveness.

## Author Contributions

SJA conceived and designed the study, conducted the statistical analyses, and took the lead in drafting the manuscript. PVS and SJA were responsible for data acquisition and curation. MB and AJW contributed to the study design and concept development. AJW, BAH, PVS, and MB provided clinical and scientific guidance and contributed to the interpretation of the results. All authors critically reviewed, revised, and approved the final version of the manuscript.

## Abbreviations

T2D: type 2 diabetes
VINT: Virta Individualized Nutrition Therapy
UC: usual care
LRE: liver related event
MASLD: metabolic dysfunction–associated steatotic liver disease
MASH: metabolic dysfunction–associated steatohepatitis
PHTN: portal hypertension
HCC: hepatocellular carcinoma
BHB: beta hydroxybutyrate

## Data Availability

The datasets used in this study are not publicly available due to licensing restrictions. They were obtained under a commercial agreement with the data vendor and may only be accessed in accordance with the terms of that license.

## Compliance with Ethics Guidelines

This study was deemed exempt from institutional review board (IRB) oversight because it utilized only retrospective, de-identified data and did not involve any direct interaction or intervention with human participants.

## References

1. Xia Y, Zhang L, et al. Prevalence of metabolic dysfunction-associated fatty liver disease in the United States: NHANES 2017–2020 analysis. J Hepatol. 2025;73(2):450–459. doi:10.1016/j.jhep.2025.01.015

2. Younossi ZM, Stepanova M, et al. Epidemiology of nonalcoholic fatty liver disease and nonalcoholic steatohepatitis in adults with type 2 diabetes in the United States. Diabetes Care. 2025;48(3):555–564. doi:10.2337/dc24-1984

3. Huang DQ, Wong VWS, Rinella ME, Boursier J, Lazarus JV, Yki-Järvinen H, Loomba R. Metabolic dysfunction–associated steatotic liver disease in adults. Nat Rev Dis Primers. 2025;11:14. doi:10.1038/s41572-025-00599-1.

4. Lazarus JV, Mark HE, Anstee QM, Arab JP, Batterham RL, Castera L, et al.; NAFLD Consensus Consortium. Advancing the global public-health agenda for NAFLD: a consensus statement. Nat Rev Gastroenterol Hepatol. 2022;19:60–78. doi:10.1038/s41575-021-00523-4.

5. Ciardullo S, Perseghin G. Prevalence of NAFLD and advanced fibrosis in overweight and obese US adults: NHANES elastography study. Liver Int. 2024;44(3):512–520. doi:10.1111/liv.15632

6. Estes C, Anstee QM, et al. Modeling the burden of NASH and advanced fibrosis in the United States using NHANES. Hepatol Commun. 2024;8(5):713–724. doi:10.1002/hep4.237

7. Younossi ZM, Blachier M, Li C, et al. Projected global clinical, humanistic, and economic impact of metabolic dysfunction–associated steatohepatitis (MASH): the cost of inaction. Clin Gastroenterol Hepatol. 2025; doi:10.1016/j.cgh.2025.09.002.

8. Estes C, Razavi H, Loomba R, Younossi Z, Sanyal AJ. Modeling the epidemic of nonalcoholic fatty liver disease and its impact on advanced liver disease and mortality in the United States. Hepatology. 2018;67(1):123–133. doi:10.1002/hep.29466

9. Vilar-Gomez E, Martinez-Perez Y, Calzadilla-Bertot L, et al. Weight loss through lifestyle modification significantly reduces features of nonalcoholic steatohepatitis. Gastroenterology. 2015;149(2):367–378.e5. doi:10.1053/j.gastro.2015.04.005

10. Jensen M, Koppe S, Andersen TW, et al. A narrative review of lifestyle management in MASLD/MASH. Hepatology. 2024;80(3):615–632. doi:10.1002/hep.998.

11. Chalasani N, Younossi Z, Lavine JE, et al. The diagnosis and management of nonalcoholic fatty liver disease: Practice guidance from the American Association for the Study of Liver Diseases. Hepatology. 2018;67(1):328–357. doi:10.1002/hep.29367

12. Rinella ME, Neuschwander-Tetri BA, Siddiqui MS, et al. AASLD Practice Guidance on the clinical assessment and management of nonalcoholic fatty liver disease. Hepatology. 2023;77(5):1797–1835. doi:10.1097/HEP.0000000000000323

13. European Association for the Study of the Liver (EASL), European Association for the Study of Diabetes (EASD), European Association for the Study of Obesity (EASO). EASL–EASD–EASO Clinical Practice Guidelines for the management of non-alcoholic fatty liver disease. J Hepatol. 2016;64(6):1388–1402. doi:10.1016/j.jhep.2015.11.004

14. Harrison SA, Bedossa P, Guy CD, Schattenberg JM, Loomba R, et al; MAESTRO-NASH Investigators. A phase 3, randomized, controlled trial of resmetirom in NASH with liver fibrosis. N Engl J Med. 2024;390(6):497–509. doi:10.1056/NEJMoa2309000

15. Sanyal AJ, Newsome PN, Kliers I, et al; ESSENCE Study Group. Phase 3 trial of semaglutide in metabolic dysfunction-associated steatohepatitis. N Engl J Med. 2025;392(21):2089–2099. doi:10.1056/NEJMoa2413258

16. Hallberg SJ, McKenzie AL, Williams PT, et al. Effectiveness and safety of a novel care model for the management of type 2 diabetes at 1 year: an open-label, nonrandomized, controlled study. Diabetes Ther. 2018;9(2):583–612. doi:10.1007/s13300-018-0373-9

17. Athinarayanan SJ, Adams RN, Hallberg SJ, et al. Long-term effects of a novel continuous remote care intervention including nutritional ketosis for the management of type 2 diabetes: a 2-year nonrandomized clinical trial. Front Endocrinol (Lausanne). 2019;10:348. doi:10.3389/fendo.2019.00348

18. McKenzie AL, Athinarayanan SJ, McCue J, et al. Type 2 diabetes prevention focused on normalization of glycemia: a two-year pilot study. Nutrients. 2021;13(3):749. doi:10.3390/nu13030749

19. Roberts CGP, Athinarayanan SJ, Volk BM, Adams RN. One-year real-world outcomes of remotely delivered carbohydrate-restricted nutrition therapy. Obesity (Silver Spring). 2024;32(1):55–308. doi:10.1002/oby.24195

20. Athinarayanan SJ, Shanmugam PV, Roberts CGP. Cost-effective long-term weight loss and maintenance via nutritional ketosis and telemedicine, without GLP1ra. [embargoed].

21. Vilar-Gomez E, Athinarayanan SJ, Adams RN, et al. Post hoc analyses of surrogate markers of nonalcoholic fatty liver disease and liver fibrosis in patients with type 2 diabetes in a digitally supported continuous care intervention: an open-label, nonrandomized controlled study. BMJ Open. 2019;9(3):e023597. doi:10.1136/bmjopen-2018-023597

22. Khan T, Kim C, Druet A. High burden, low priority? Iron deficiency anemia in U.S. women and the challenges of data gaps with over-the-counter treatments. Komodo Insights 9 Oct 2023. https://www.komodohealth.com/insights/high-burden-low-priority-iron-deficiency-anemia-in-u.s-women-and-the-challenges-of-data-gaps-with-over-the-counter-treatment (accessed 15 Oct 2025).

23. von Elm E, Altman DG, Egger M, Pocock SJ, Gøtzsche PC, Vandenbroucke JP; STROBE Initiative.The Strengthening the Reporting of Observational Studies in Epidemiology (STROBE) statement: guidelines for reporting observational studies. BMJ. 2007;335(7624):806–808. doi:10.1136/bmj.39335.541782.AD

24. Abeysekera KWM, Valenti L, Younossi Z, et al. Implementation of a liver health check in people with type 2 diabetes. Lancet Gastroenterol Hepatol. 2024;9(1):83–91. doi:10.1016/S2468-1253(23)00270-4.

25. Lazarus JV, Agirre-Garrido L, Miralles-Sánchez JE, et al,. Cost of Metabolic Dysfunction-Associated Steatotic Liver Disease Screening Among All People Living With Diabetes in Six Countries. Liver Int. 2025;45:e70390. doi:10.1111/liv.70390.

26. Lazarus JV, Agirre-Garrido L, Díaz LA, et al. Cost-effectiveness of MASH diagnosis and management approaches among those with type 2 diabetes. JAMA Netw Open. 2025;8(11):e2542750. doi:10.1001/jamanetworkopen.2025.42750.

27. Ivancovsky-Wajcman D, Brennan PN, Kopka CJ, et al. Integrating social nutrition principles into the treatment of steatotic liver disease. Commun Med. 2023;3:165. doi:10.1038/s43856-023-00398-3.

28. Zelber-Sagi S, Carrieri P, Pericàs JM, et al. Food inequity and insecurity and MASLD: burden, challenges, and interventions. Nat Rev Gastroenterol Hepatol. 2024;21(3):668–686. doi:10.1038/s41575-024-00959-4.

29. Kanwal F, Kramer JR, Li L, et al. GLP-1 receptor agonists and risk for cirrhosis and related complications in patients with metabolic dysfunction–associated steatotic liver disease. JAMA Intern Med. 2024;184(11):1314–1323. doi:10.1001/jamainternmed.2024.4661

30. Jung C, Park S, Kim H. Association between hypoglycemic agent use and the risk of occurrence of nonalcoholic fatty liver disease in patients with type 2 diabetes mellitus. PLoS One. 2023;18(11):e0294423. doi:10.1371/journal.pone.0294423

31. Dulai PS, Singh S, Patel J, et al. Increased risk of mortality by fibrosis stage in nonalcoholic fatty liver disease: systematic review and meta-analysis. Hepatology. 2017;65(5):1557–1565.

32. Younossi ZM, Anstee QM, Marietti M, et al. Global burden of NAFLD and NASH: trends, predictions, risk factors, and prevention. Nat Rev Gastroenterol Hepatol. 2018;15(1):11–20. doi:10.1038/nrgastro.2017.109

33. Niu J, Al-Yaman W, Pinyopornpanish K, et al. The long-term effect of weight loss on the prevention of progression to cirrhosis among patients with obesity and MASH-related F3 liver fibrosis. Int J Environ Res Public Health. 2024;21(6):708. doi:10.3390/ijerph21060708

34. Perreault L, Kramer ES, Smith PC, et al. A closer look at weight loss interventions in primary care: a systematic review and meta-analysis. Front Med (Lausanne). 2023;10:1204849. doi:10.3389/fmed.2023.1204849

35. Pagoto S, et al. An evidence-based guide for obesity treatment in primary care. Am J Med. 2015;128(10):e11–e19. doi:10.1016/j.amjmed.2015.07.015

36. Oshman L, Othman A, Furst W, et al. Primary care providers’ perceived barriers to obesity treatment and opportunities for improvement: a mixed methods study. PLoS One. 2023;18(4):e0284474. doi:10.1371/journal.pone.0284474

37. Dudekula A, Rachakonda V, Shaik B, Behari J. Weight loss in nonalcoholic fatty liver disease: clinical outcomes and predictors of success. PLoS One. 2014;9(11):e111808. doi:10.1371/journal.pone.0111808

38. Hallsworth K, Adams LA. Lifestyle modification in NAFLD/NASH: facts and figures. JHEP Rep. 2019;1(6):468–479. doi:10.1016/j.jhepr.2019.10.008

39. McKenzie AL, Athinarayanan SJ, Van Tieghem MR, et al. Five-year effects of a novel continuous remote care model with carbohydrate-restricted nutrition therapy including nutritional ketosis in type 2 diabetes: an extension study. Diabetes Res Clin Pract. 2024;217:111898. doi:10.1016/j.diabres.2024.111898

40. Adams RN, Athinarayanan SJ, Zoller AR, McKenzie AL, Ratner RE. Sustained metabolic improvements in a remotely delivered ketogenic nutrition program for veterans with type 2 diabetes: a 3-year observational study. Diabetes Obes Metab. 2025;27(1):e16525. doi:10.1111/dom.16525

41. McKenzie AL, Athinarayanan SJ. Impact of glucagon-like peptide-1 agonist deprescription in type 2 diabetes in a real-world setting: a propensity score–matched cohort study. Diabetes Ther. 2024;15(10):1–11. doi:10.1007/s13300-024-01547-0

42. Finucane FM, Rafey MF, Leahy M, et al. Weight loss is proportional to increases in fasting serum β-hydroxybutyrate concentrations in adults with severe obesity undergoing a low-energy meal replacement program. Hum Nutr Metab. 2023;200192. doi:10.1016/j.hnm.2023.200192

43. McKenzie AL, Athinarayanan SJ, Adams RN, et al. Mean blood β-hydroxybutyrate predicts clinically significant weight loss following 90 days of carbohydrate-restricted nutrition therapy. Diabetes. 2021;70(suppl 1):307-OR. doi:10.2337/db21-307-OR

44. Adams RN, Athinarayanan SJ, Roberts CGP, Volk BM. Real-world ketone targets in carbohydrate-restricted nutrition therapy for obesity. Obesity (Silver Spring). 2024;32(1):55–308. doi:10.1002/oby.24195

45. Youm YH, Nguyen KY, Grant RW, et al. The ketone metabolite β-hydroxybutyrate blocks NLRP3 inflammasome–mediated inflammatory disease. Nat Med. 2015;21(3):263–269. doi:10.1038/nm.3804

46. Goldberg EL, Molony RD, Kudo E, et al. Ketogenesis activates metabolically protective γδ T cells in visceral adipose tissue. Cell Metab. 2020;31(3):420–436.e7. doi:10.1016/j.cmet.2020.01.009

47. Mardinoglu A, Wu H, Bjornson E, et al. An integrated understanding of the rapid metabolic benefits of a carbohydrate-restricted diet on hepatic steatosis in humans. Cell Metab. 2018;27(3):559–571.e5. doi:10.1016/j.cmet.2018.01.005

48. Hyde PN, Newsom SA, Sapper TN, et al. Dietary carbohydrate restriction improves metabolic syndrome independent of weight loss. JCI Insight. 2019;4(12):e128308. doi:10.1172/jci.insight.128308

49. Shimazu T, Hirschey MD, Newman J, et al. Suppression of oxidative stress by β-hydroxybutyrate, an endogenous histone deacetylase inhibitor. Science. 2013;339(6116):211–214. doi:10.1126/science.1227166

50. Puchalska P, Crawford PA. Multi-dimensional roles of ketone bodies in fuel metabolism, signaling, and therapeutics. Cell Metab. 2017;25(2):262–284. doi:10.1016/j.cmet.2016.12.022

51. Harrison SA, Taub R, Neff GW, et al. Resmetirom for nonalcoholic fatty liver disease: a randomized, double-blind, placebo-controlled phase 3 trial. Nat Med. 2023;29(11):2919–2928. doi:10.1038/s41591-023-02603-1

52. Effect of Resmetirom or Placebo in NASH Fibrosis Patients With <5% or >5% Weight Loss and/or on Baseline GLP-1 Therapy in the MAESTRO-NASH 52-Week Serial Liver Biopsy Study. Gastroenterol Hepatol (N Y). 2024;20(12 Suppl 11):8–9.

53. Zhou XD, Wong VWS, Zheng MH. Resmetirom and GLP-1 agonists for MASH: complementary rather than exclusive. NPJ Gut Liver. 2024;1:4. doi:10.1038/s44355-024-00004-w

54. Lazarus JV, Ivancovsky Wajcman D, Mark HE, et al. Opportunities and challenges following approval of resmetirom for MASH liver disease. Nat Med. 2024;30:3402–3405. doi:10.1038/s41591-024-02958-z.

55. U.S. Food and Drug Administration (FDA). Noncirrhotic Nonalcoholic Steatohepatitis With Liver Fibrosis: Developing Drugs for Treatment—Guidance for Industry. Silver Spring, MD: U.S. Department of Health and Human Services, Food and Drug Administration; 2018. Available at: https://www.fda.gov/media/119044/download

