## Supplemental Material for "Reduced Risk of Liver Related Events Among Patients Receiving Individualized Nutrition-Focused Remote Care in the United States"

**Supplementary Method**

**Virta Program Eligibility**

Adults aged 18 years and older with overweight, obesity, prediabetes, or type 2 diabetes (T2D) were eligible for enrollment in the individualized nutrition therapy program designed for diabetes reversal or sustainable weight management. Participants were required to have the capacity to engage with the digital care platform and to remain enrolled for at least one year following the index date.

Individuals were excluded if they had medical conditions that could make nutritional ketosis unsafe. These included type 1 or secondary diabetes, BMI <18.5 kg/m², ongoing cancer treatment, end-stage renal disease or advanced chronic kidney disease (stage IV–V; estimated glomerular filtration rate <30 mL/min/1.73 m²), decompensated liver disease (Child-Pugh class C), uncontrolled electrolyte disturbances, severe hypertriglyceridemia (triglycerides >4000 mg/dL), pancreatic insufficiency or significant malabsorption, active eating disorders, alcohol misuse, recurrent arrhythmias or unexplained syncope, New York Heart Association (NYHA) class IV heart failure, neurologic or psychiatric illness impairing self-management, contraindicated medications (e.g., desmopressin, tolvaptan), pregnancy or exclusive breastfeeding, rare metabolic disorders, or other conditions deemed unsafe by the clinical team.

Certain conditions required individualized review prior to enrollment, including BMI 18.5–20 kg/m², moderate alcohol use (>2 drinks/day with willingness to reduce), NYHA class III heart failure, CKD stage IIIB, Child-Pugh class B liver disease, latent autoimmune diabetes in adults (LADA), history of eating disorders in remission, partial pancreatectomy, moderate hypertriglyceridemia (2000–4000 mg/dL), chronic steroid or immunosuppressant use, lithium therapy, cognitive impairment, or breastfeeding during weaning. Individuals with stable chronic conditions such as well-controlled arrhythmias, CKD stage I–IIIA, Child-Pugh class A liver disease, and stable psychiatric disorders (e.g., depression, bipolar disorder, anxiety) were considered eligible.

##### **Virta Nutritional and Lifestyle Intervention**

Participants received a personalized nutritional intervention centered on carbohydrate restriction to achieve and maintain nutritional ketosis, with the goal of improving glycemic and cardiometabolic health. The target β-hydroxybutyrate (BHB) range was 0.5–3.0 mmol/L, consistent with physiologic nutritional ketosis. Protein intake was maintained at moderate levels (approximately 1.2–1.5 g/kg reference body weight per day), while dietary fat served as the primary energy source, adjusted to satiety and individual energy needs. The dietary pattern emphasized whole, unprocessed foods such as non-starchy vegetables, healthy fats (e.g., olive oil, nuts, avocado), and moderate portions of protein from meat, fish, poultry, eggs, and dairy, while minimizing refined carbohydrates, added sugars, and starch-rich foods.

Participants received structured education on hydration, sodium and electrolyte replacement, and management of transient adaptation symptoms (e.g., “keto flu”). The intervention was delivered through a telemedicine platform that provided continuous access to licensed medical providers and health coaches. Participants were supported through one-on-one and group-based digital education sessions addressing nutrition, behavior change, and lifestyle adaptation.

##### **Biomarker Monitoring**

Monitoring of β-hydroxybutyrate (BHB) was a core element of both safety assessment and adherence tracking. Participants were provided with FDA-cleared, at-home blood ketone meters capable of measuring both glucose and BHB using fingerstick samples. During the initial phase of carbohydrate restriction (typically the first 2–4 weeks), participants were instructed to log daily BHB values to monitor metabolic adaptation and identify early deviations from nutritional ketosis.

Once stable ketosis was achieved, the frequency of BHB logging transitioned to a weekly schedule and was further individualized based on participant preferences, metabolic stability, medication use, and provider recommendations. Participants undergoing medication adjustments, those with variable glycemic control, or those requiring additional dietary support were encouraged to monitor more frequently.

All BHB values were transmitted automatically via connected devices or manually entered into the digital application and reviewed by the care team in real time. Providers monitored BHB trends longitudinally to evaluate adherence, assess metabolic response, and guide ongoing nutrition and medication adjustments. Persistently low BHB values triggered additional coaching and dietary reinforcement, whereas sustained readings above 3.0 mmol/L prompted review for potential dehydration, caloric insufficiency, or intercurrent illness.

In addition to BHB, participants tracked blood glucose, blood pressure, and body weight. These biomarkers were recorded daily during program initiation, particularly within the first three months, and subsequently adjusted to a participant-specific schedule reflecting their clinical stability. Continuous integration of these data enabled proactive provider feedback, early identification of safety concerns, and dynamic care plan adjustments to support long-term adherence and success.

##### **Medication Management**

A structured, provider-led medication management protocol ensured participant safety during carbohydrate restriction and facilitated medication optimization as metabolic health improved. At program initiation, clinicians reviewed each participant’s medication profile and developed individualized tapering plans. Medications associated with high hypoglycemia risk, such as insulin and sulfonylureas, were typically reduced or discontinued early in the intervention. Antihypertensive agents were adjusted in response to improved blood pressure, and diuretics were reassessed to prevent dehydration and electrolyte disturbances due to increased natriuresis.

Other cardiometabolic therapies including metformin, GLP-1 receptor agonists, SGLT2 inhibitors, and lipid-lowering agents were modified based on clinical response, comorbidities, and laboratory findings. If participants were unable to sustain carbohydrate restriction or demonstrated worsening glycemia, medications were reintroduced as clinically indicated.

Daily participant-reported data, including BHB, glucose, blood pressure, and weight, were reviewed continuously by the remote care team through the digital platform. This proactive and adaptive approach balanced deprescription with clinical safety, minimizing risks such as hypoglycemia and hypotension, while allowing for evaluation of the independent effects of nutritional ketosis on glycemic control, blood pressure, and other cardiometabolic outcomes.

**Statistical Method**

#### **Supplementary Statistical Methods**

##### **Cohort assembly and baseline exclusions**

Analyses were performed in Komodo Health’s Sentinel environment using deidentified claims and enrollment data from the Healthcare Map™. Before any balancing, we excluded persons with baseline evidence of advanced or chronic liver disease to define an at-risk cohort. Exclusions included cirrhosis, portal hypertension and its complications (ascites, varices, hepatic encephalopathy), hepatocellular carcinoma, alcohol associated liver disease, viral hepatitis, and autoimmune or metabolic liver disorders. Individuals with MASLD, MASH, or fibrosis at baseline were retained so that incident and progression outcomes could be evaluated within these higher risk strata.

##### **Baseline variables and summaries**

Continuous variables were summarized as mean (SD) and categorical variables as n (%). Between group balance was described using standardized mean differences (SMD), with absolute SMD ≤ 0.10 indicating adequate balance. No hypothesis tests were performed for baseline characteristics.

Baseline covariates for modeling and balancing included demographics (age in coarse categories with a missingness indicator, sex, race or ethnicity), clinical conditions (type 2 diabetes, obesity, hypertension, hyperlipidemia, coronary artery disease, stroke, heart failure, chronic kidney disease stage, albuminuria, smoking, alcohol related diagnoses, and other cardiovascular disease), liver condition flags (baseline MASLD, MASH, fibrosis), baseline liver procedure indicator, and medication exposures at baseline (SGLT2 inhibitors, GLP1 receptor agonists, statins, insulin, sulfonylureas, DPP4 inhibitors, other type 2 diabetes therapies and medications with potential liver benefits including certain statins, icosapent ethyl, omega-3 acid ethyl esters, Vitamin E, alpha-tocopherol, gemfibrozil and fenofibrate). One-year pre index utilization and costs were included as continuous covariates for inpatient, outpatient, and prescription spending, as well as total cost of care. Census region and area deprivation index quintile were added to account for geographic and socioeconomic variation.

##### **Propensity score matching**

We estimated the probability of enrollment in the Virta Individualized Nutrition Therapy program vs usual care using logistic regression with the covariates listed above. Matching used the MatchIt package in R with nearest neighbor 1 to 1 matching without replacement on the logit of the propensity score.

- Distance: logit of the estimated propensity score
- Ratio: 1
- Replacement: false
- Exact matching was enforced on sex, age category, race or ethnicity, and baseline liver condition indicators for MASLD, MASH, and fibrosis, drugs with potential liver benefits including incretin mimetics and SGLT2i
- Balance after matching was assessed with SMD and Love plots using the cobalt package. A threshold of 0.10 was used for acceptable balance.

##### **Inverse probability of treatment weighting**

As a complementary approach, inverse probability of treatment weighting was conducted using the same covariate set as the matching model. Stabilized weights were computed. Participants with extreme propensity scores were trimmed to improve overlap. Weighted Cox proportional hazards models with robust variance estimators were fit in the weighted sample. Balance after weighting was reevaluated with SMD and the 0.10 threshold.

##### **Outcomes and follow up**

Primary composite outcomes are defined in Study Outcomes. Time zero was the index date. Participants were followed until the first qualifying outcome, disenrollment, or end of study, with a maximum of 5 years.

##### **Primary time to event analyses**

Treatment effects were estimated using Cox proportional hazards models fit to the matched cohort and stratified by matched set. Robust standard errors were clustered on matched pairs. Base models adjusted for age, sex, and race or ethnicity. Extended models additionally adjusted for baseline type 2 diabetes and obesity and for follow up exposure to cardiometabolic medications with plausible hepatic effects. Medication use was represented in two ways: ever or never exposure during follow up, and proportion of days covered to reflect adherence for drugs or supplements with liver benefits (potential MASLD), SGLT2 inhibitors, GLP1 receptor agonists, TZD when available, and statins. Proportional hazards assumptions were checked using Schoenfeld residuals and visual diagnostics.

Incidence rates were calculated as events per 1,000 person years. Kaplan Meier curves were produced with numbers at risk. Absolute risk reduction was computed as the difference in incidence rates between usual care and VINT, and the number needed to treat was calculated as 1 divided by absolute risk reduction.

##### **Secondary analyses within the VINT cohort**

Associations between clinical response markers and the primary composite outcome were estimated with separate Cox models for each categorical predictor:

1. mean beta hydroxybutyrate ≥ 0.3 vs < 0.3 mmol per L with missing grouped with < 0.3
2. mean beta hydroxybutyrate ≥ 0.5 vs < 0.5 mmol per L with missing grouped with < 0.5
3. 6-month weight change ≥ 10% vs < 10%
4. 6-month weight change ≥ 15% vs < 15%
5. HbA1c change ≥ 0.5 percentage points vs < 0.5 points using 1 year values when available or 6-month values otherwise

When indicated, models were further adjusted for follow up medication exposure using the ever or never and proportion of days covered specifications described above. Incidence rates were compared across weight-loss categories using Poisson regression to estimate rate ratios and 95% confidence intervals, with the <5% weight-loss group as reference. For prevalence comparisons, Chi-square and Cochran–Armitage trend tests were used to assess differences and linear trends across increasing weight-loss categories.

Sensitivity analysis

Participants who developed other liver diseases (e.g., alcohol associated liver disease, viral hepatitis, or autoimmune hepatitis) during follow up were excluded, and advanced liver outcomes were reassessed within this restricted cohort. A second analysis adjusted for healthcare utilization variables, including outpatient costs, average number of imaging studies, laboratory tests, and clinical evaluations per member per year, to account for potential differences in healthcare contact or diagnostic intensity. Because death may act as a competing event for liver outcomes, a Fine Gray sub distribution hazards model was applied, specifying death from any cause as a competing risk for advanced liver disease. Finally, all-cause mortality was evaluated using a Cox proportional hazards model to complement the competing risk analysis.

##### **Exploratory analyses**

Exploratory analyses included a subcohort of participants with baseline MASLD, MASH, or fibrosis and at least one follow-up diagnosis recorded on a new date. Transitions between liver disease states were evaluated descriptively. Liver-specific imaging, procedures, and biopsy were assessed descriptively in the matched cohort, with overall distributions compared using Wilcoxon rank-sum tests. Medication use was evaluated across predefined metabolic and liver-directed therapy classes (e.g., GLP-1 receptor agonists, SGLT2 inhibitors, thiazolidinediones [pioglitazone], vitamin E, statins, and other supplements with liver benefits). The proportion of participants with any exposure and the cumulative duration of use (mean and median days covered) were summarized, and between-group differences were assessed to contextualize treatment patterns.

##### **Software**

All analyses were conducted in R version 4.3.1 within Komodo Health’s Sentinel environment. Core packages included survival, survminer, MatchIt, cobalt, tableone, gtsummary, broom, ggplot2, sandwich, dplyr, tidyverse, lubridate, tidyr, stringr, reshape2, and purrr. Model outputs were tidied with broom and exported as CSV files for documentation.

**Supplementary Results**

**Fine Gray Subdistribution Model and Mortality Outcome**

Mortality information was derived from multiple linked data sources; however, the cause of death was not available and therefore was not used as an endpoint in the liver-related events analysis. Because specific causes of death could not be determined, all-cause mortality was included solely as a sensitivity analysis to assess the potential impact of competing risk. Using mortality data restricted to deaths occurring within the closed claims coverage period to ensure completeness, a Fine Gray subdistribution hazards model was performed, specifying death from any cause as a competing event for the primary liver outcome of advanced liver disease across analytic strategies 1 through 3. In this model, the cumulative incidence of advanced liver disease remained significantly lower among participants in the VINT group compared with controls (SHR 0.27, 95% CI 0.16–0.44, *p* < 0.001). This finding was consistent for the advanced liver disease outcome across all three analytic strategies. In addition, all-cause mortality was evaluated using a Cox proportional hazards model, which also demonstrated a lower risk of death in the VINT group (1.0 vs. 3.8 per 1,000 person years; HR 0.25, 95% CI 0.12–0.50, *p* < 0.001). Given that overall mortality was lower among VINT participants, any competing risk bias would be expected to attenuate rather than exaggerate the difference in liver event incidence between groups. These results further support the robustness of the primary findings.

**Secondary outcomes**

In the VINT cohort, weight loss of ≥15% was significantly associated with a reduced risk of new onset liver disease in demographic-adjusted models (incidence rate 21.2 per 1,000 person-years in ≥15% vs 31.8 per 1,000 person-years in <15%; HR = 0.66, 95% CI: 0.46–0.95, p = 0.02). In contrast, weight loss of ≥10% was marginally significantly associated with risk reduction (HR = 0.78, 95% CI: 0.61–1.00, p = 0.05). After further adjustment for medication use, the association for ≥15% weight loss was attenuated (HR = 0.67, 95% CI: 0.46–0.97, p = 0.04 for categorical medications; HR = 0.73, 95% CI: 0.51–1.04, p = 0.08 for continuous PDC medications). A dose–response relationship was observed across weight-loss categories, with lower incidence (Figure 4A) and prevalence of liver disease at greater weight reduction (Figures 4B). Only the ≥15 % group showed a significantly lower incidence rate versus <5 % (*p* = 0.01), and the overall trend in prevalence was significant (*p* < 0.001). Mean BHB levels over 6 months were not significantly associated with incident liver disease: ≥0.5 mmol/L (HR = 0.88, 95% CI: 0.69–1.12, p = 0.30) and ≥0.3 mmol/L (HR = 1.08, 95% CI: 0.81–1.44, p = 0.58). Similarly, HbA1c reduction was not significantly associated with risk of new onset liver disease (HR = 1.20, 95% CI: 0.94–1.55, p = 0.14)

**Exploratory outcomes**

**Liver specific imaging, procedures and medication persistence**

In the analysis of liver specific imaging and biopsy procedures within the matched cohort, utilization was consistently lower among participants in the VINT program compared with UC. Across individual imaging modalities such as abdominal ultrasound, CT, MRI, and elastography (FibroScan), absolute differences were modest; however, when all liver related diagnostic procedures were combined, the overall distribution differed significantly between groups (Wilcoxon rank sum test, *p* = 0.02; Supplementary Table S7). To further investigate this, we compared mean imaging counts per person year between the VINT and control cohorts at both 1 and 2 years follow up. The average number of imaging studies did not differ significantly between groups (mean 2.11 vs 2.01 at 1 year, *p* = 0.18; mean 3.10 vs 3.29 at 2 years, *p* = 0.08), indicating that VINT participants did not undergo fewer overall imaging tests. Similarly, evaluation counts were comparable (mean 6.11 vs 6.67 at 1 year, *p* = 0.45; mean 10.13 vs 10.83 at 2 years, *p* = 0.02), supporting that general healthcare contact remained balanced between groups. These findings suggest that the lower number of liver specific procedures observed in the VINT group is unlikely to reflect reduced testing intensity but likely fewer clinical indications for imaging, consistent with improved metabolic and liver health.

Medication use patterns were broadly similar between groups (Supplementary Table S8), and resmetirom use remained rare (0.3% vs 0.1%).

**Supplementary Discussion**

**Limitation of the Study**

Several limitations should be acknowledged. Both baseline exclusions and incident MASLD outcomes were identified using ICD 10 codes, as anthropometric, laboratory, and imaging data were unavailable in the external control cohort. The Komodo database is a claims-based dataset without linked electronic health record data; therefore, variables such as body weight, BMI, A1c, and lipid profiles could not be incorporated into matching or outcome ascertainment. Consequently, some individuals with undiagnosed MASLD or MASH may not have been captured, introducing potential under ascertainment or misclassification. The requirement for one year of continuous medical and pharmacy coverage ensured data completeness but may have preferentially included individuals with more stable insurance and stronger healthcare engagement; only 10–15 percent of individuals in large claims datasets typically meet strict continuous enrollment criteria. Participants who remained in the VINT program for at least one year may represent a more adherent population, although this threshold ensures adequate exposure and follow up. The observation of fewer liver imaging procedures among VINT participants may reflect improved metabolic health rather than reduced testing frequency. This interpretation is supported by the healthcare utilization sensitivity analysis, the balanced medication uses between groups, and the finding that overall healthcare contact including all imaging, not only liver-related imaging as well as evaluation counts during follow-up were similar in both groups. Mortality data were restricted to deaths captured within the closed claims coverage window, and cause of death was unavailable; therefore, all-cause mortality was used solely as a sensitivity analysis. The relatively short mean follow up may have limited the ability to observe late-stage hepatic complications. Ketone exposure was assessed only during the first six months, which may not reflect sustained nutritional ketosis. Finally, the study population was predominantly commercially insured, although we do not have income data, the 2 highest ADI quintiles were almost identical in both groups. This may somewhat limit generalizability to lower income or more diverse populations, and residual confounding from unmeasured factors cannot be excluded.

### **STROBE Statement—Checklist of Items That Should Be Included in Reports of Cohort Studies**

This checklist has been completed for the manuscript titled “Reduced Onset of MASLD, MASH, and Advanced Liver Disease in Patients Who Received Individualized Nutrition-Focused Remote Care for Adults with Type 2 Diabetes and Obesity,” prepared for submission to Hepatology. Each item of the STROBE statement is addressed below, with corresponding page numbers and section locations in the manuscript and supplementary materials.

| **Item No.** | **STROBE Checklist Item** | **Location in Manuscript and Supplementary Materials** |
| --- | --- | --- |
| **1** | Title and abstract: Indicate the study’s design in the title or abstract; provide an informative summary. | Title page, Abstract (pp. 1–2) |
| **2** | Background/rationale: Explain the scientific background and rationale for the investigation being reported. | Introduction (pp. 3–5) |
| **3** | Objectives: State specific objectives, including any prespecified hypotheses. | Introduction, final paragraph (p. 5) |
| **4** | Study design: Present key elements of the study design early in the paper. | Methods, Data Source and Study Design (pp. 6–7) |
| **5** | Setting: Describe the setting, locations, and relevant dates, including periods of recruitment, exposure, follow-up, and data collection. | Methods, Data Source (pp. 6–7); Supplementary Methods, Data Source (pp. 1–2) |
| **6** | Participants: Give eligibility criteria, sources and methods of selection, and methods of follow-up. | Methods, Study Population and Intervention (pp. 7–8); Supplementary Methods, Eligibility Criteria (pp. 1–2) |
| **7** | Variables: Clearly define all outcomes, exposures, predictors, potential confounders, and effect modifiers; give diagnostic criteria, if applicable. | Methods, Study Outcomes (pp. 8–9) |
| **8** | Data sources/measurement: For each variable of interest, give sources of data and methods of assessment (measurement). | Methods, Data Source (pp. 6–7); Supplementary Methods, Biomarker Monitoring (pp. 3–4) |
| **9** | Bias: Describe any efforts to address potential sources of bias. | Methods, Statistical Analysis and Sensitivity Analyses (pp. 9–10); Supplementary Methods, Statistical Method (pp. 5–7) |
| **10** | Study size: Explain how the study size was arrived at. | Methods, Study Population and Matching (p. 7); Supplementary Methods, Cohort Assembly (p. 5) |
| **11** | Quantitative variables: Explain how quantitative variables were handled in the analyses. Describe groupings and rationale. | Methods, Statistical Analysis (pp. 9–10); Supplementary Methods, Statistical Analyses (pp. 6–8) |
| **12** | Statistical methods: Describe all statistical methods, including confounder control, subgroup analyses, missing data handling, and sensitivity analyses. | Methods, Statistical Analysis (pp. 9–10); Supplementary Methods, Statistical Methods (pp. 5–8) |
| **13** | Participants: Report numbers of individuals at each study stage (e.g., eligible, included, followed up, analyzed). | Results, Study Participant Flow (pp. 10–11); Figure 1 (p. 23) |
| **14** | Descriptive data: Give characteristics of study participants and information on exposures and potential confounders. | Results, Baseline Characteristics (pp. 11–12); Tables 1–2 (pp. 24–26) |
| **15** | Outcome data: Report numbers of outcome events or summary measures over time. | Results, Primary Outcomes (pp. 12–14); Tables 3–4 (pp. 27–28) |
| **16** | Main results: Give unadjusted and adjusted estimates with 95% confidence intervals; clarify confounders adjusted for and rationale. | Results, Primary and Secondary Outcomes (pp. 12–15); Tables 3–4 (pp. 27–28) |
| **17** | Other analyses: Report additional analyses such as subgroup, sensitivity, and exploratory analyses. | Results, Sensitivity and Exploratory Analyses (pp. 15–16); Supplementary Tables S5–S8 (pp. 10–12) |
| **18** | Key results: Summarize key results with reference to study objectives. | Discussion, opening paragraph (pp. 17–18) |
| **19** | Limitations: Discuss study limitations, addressing potential sources of bias or imprecision. | Discussion, limitations section (p. 21) |
| **20** | Interpretation: Provide an overall interpretation considering objectives, limitations, multiplicity of analyses, results from similar studies, and other evidence. | Discussion, interpretation (pp. 18–22) |
| **21** | Generalisability: Discuss the generalisability (external validity) of the study results. | Discussion, final paragraphs (pp. 22–23) |
| **22** | Funding: Give the source of funding and the role of the funders for the present study. | Footnotes: Financial Support and Sponsorship section (p. 2) |

**Supplementary Figure S1.** Distribution of index dates among treated participants and matched controls. Bars represent the count of participants initiating follow-up on a given index date, plotted separately for VINT (red) and UC (blue). The figure illustrates the temporal overlap in cohort enrollment, showing increasing accrual over time with peaks corresponding to more recent years. The overlap between VINT and UC participants across the study period reduces the potential for confounding by calendar time or seasonal effects.

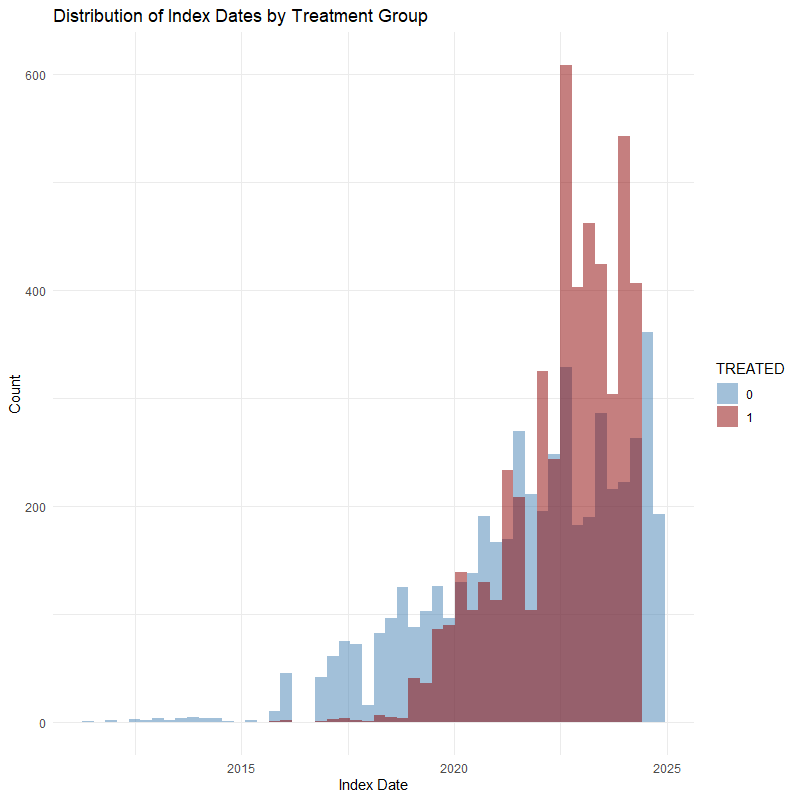

**Supplementary Table S1.** ICD-10 code sets for identifying baseline comorbidities and new-onset of all liver disease diagnoses and liver procedures during follow-up

| ICD-10 | Description |
| --- | --- |
| All liver disease diagnoses (Primary Outcomes) | |
| K76, K7685, K760 | MASLD |
| K758, K7580, K7581 | MASH |
| K740, K7400, K741, K7401, K742, K7402 | Fibrosis |
| K746, K7460, K7469 | Cirrhosis |
| K766 | Portal Hypertension |
| Portal Hypertension complications | |
| R180, R188, R189 | Ascites |
| I8500, I8501, I8510, I8511, I864 | Varices |
| K7290, K7291, K7210, K7211, K7200. K7201, G9340, G9349 | Hepatic encephalopathy |
| C220, C228 | Hepatocellular carcinoma |
| K7689 | Abnormal liver imaging |
| K769 | Unspecified liver disease |
| K7589 | Inflammatory liver disease |
| Liver procedure codes | |
| 47001 | Biopsy, with imaging |
| 47000 | Biopsy, without imaging |
| 47100 | Biopsy, wedge (intra-operative) |
| 74183 | MRI, abdomen, with or without contrast |
| 74182 | MRI, abdomen, with contrast |
| 74181 | MRI, abdomen, without contrast |
| 74160 | CT scan, abdomen, with contrast |
| 74170 | CT scan, abdomen, with or without contrast |
| 74150 | CT scan, abdomen, without contrast |
| 76700 | Ultrasound, abdomen, complete |
| 76705 | Ultrasound, abdomen, limited |
| 91200 | Liver elastography (Fibroscan) |
| 91210 | Sheer-wave elastography |
| 49082 | Paracentesis (diagnostic) |
| 49083 | Paracentesis (therapeutic) |
| 43235 | Endoscopy-variceal surveillance/therapy |
| 43239 | Endoscopy-variceal surveillance/therapy |
| 43251 | Endoscopy-variceal surveillance/therapy |
| 43255 | Endoscopy-variceal surveillance/therapy |
| 37182 | TIPS (creation) |
| 37183 | TIPS (revision) |
| 93975 | Duplex scan-portal/abdominal vessels |
| 47380 | Ablation for HCC (laser) |
| 47381 | Ablation for HCC (RF, percutaneous) |
| 47382 | Ablation for HCC (RF, Open) |
| 47379 | Unlisted laparoscopic liver procedure |
| 37243 | Transarterial embolization/chemoembolization (TACE) |
| Exclusion Liver Condition (excluded at baseline before matching) | |
| K766 | Portal Hypertension |
| R180, R188, R189 | Ascites |
| I8500, I8501, I8510, I8511, I864 | Varices |
| K7290, K7291, K7210, K7211, K7200. K7201, G9340, G9349 | Hepatic encephalopathy |
| K746, K7460, K7469 | Cirrhosis |
| C220, C228 | Hepatocellular carcinoma |
| K70 | Alcoholic Liver Disease |
| B18 | Chronic viral hepatitis |
| K753/K754 | Non-infectious inflammatory hepatitis |
| K743/K830 | Primary biliary or sclerosing cholangitis |
| E8301, E8311, E8801 | Wilson’s disease, Hemochromatosis, Alpha-1 antitrypsin deficiency |
| K71.x | Drug-induced liver injury |
| I820 | Budd-Chiari syndrome |
| Q442-Q446 | Congenital hepatic fibrosis or cystic disease |
| Z944 | Liver transplant status |
| Baseline Comorbidities (used for matching) | |
| E11 or E13 | Type 2 diabetes |
| E10 | Type 1 diabetes |
| E66 | Obesity and BMI 30+ |
| E78 | Hyperlipidemia |
| Hypertension | |
| I10 | Essential |
| I11 | Hypertensive heart disease |
| I12 | Hypertensive chronic kidney disease |
| I13 | Hypertensive heart & chronic kidney disease |
| I15.0,2,9 | Secondary hypertension |
| I16.0, 1 | Hypertensive crisis |
| Z720 | Smoking |
| C* | Any cancer |
| O* or Z3* | Pregnancy |
| Coronary Artery Disease (CAD) | |
| Ischemic Heart Disease (I20, I23, I24, I25) | |
| I24 | Acute ischemic heart disease |
| I25 | Chronic ischemic heart disease |
| I20 | Preinfarction syndrome |
| I25 | Aneurysm of coronary vessels |
| I20.1/8/9 | Angina pectoris |
| I25.81 | Arteriosclerosis of coronary artery bypass graft |
| I25.10 | Atherosclerosis of coronary artery without angina pectoris |
| I24.0 | Coronary thrombosis not resulting in myocardial infarction |
| I25.6 | Silent myocardial ischemia |
| I25.111 | Coronary artery spasm |
| I25.82 | Chronic total occlusion of coronary artery |
| I20.0 | Unstable angina co-occurrent and due to coronary arteriosclerosis |
| I25.119 | Angina co-occurrent and due to coronary arteriosclerosis |
| I25.42 | Dissection of coronary artery |
| I25.811 | Coronary arteriosclerosis in artery of transplanted heart |
| I25.812 | Arteriosclerosis of coronary artery bypass graft of transplanted heart |
| I25.1 | Coronary atherosclerosis |
| I25.719 | Arteriosclerosis of autologous vein coronary artery bypass graft |
| I25.739 | Arteriosclerosis of nonautologous coronary artery bypass graft |
| I25.729 | Arteriosclerosis of autologous arterial coronary artery bypass graft |
| I23.7 | Post infarct angina |
| I23.1 | Atrial septal defect due to and following acute myocardial infraction |
| I25.5 | Generalized ischemic myocardial dysfunction |
| I23.0,3,4,5,6 | Rupture of chordae tendinae due to and following acute myocardial infarction |
|  | Rupture of papillary muscle as current complication following acute myocardial infraction |
|  | Hemopericardium due to and following acute myocardial infraction |
|  | Rupture of cardiac wall without hemopericardium as current complication following acute myocardial infarction |
|  | Thrombosis of atrium, auricular appendage, and ventricle due to and following acute myocardial infraction |
| Heart attack (I21) | |
| I21.X | Acute non-ST segment elevation myocardial infarction |
|  | Acute ST segment elevation myocardial infraction |
|  | Myocardial infarction |
|  | Myocardial infarction due to demand ischemia |
|  | Acute ST segment elevation myocardial infarction involving left anterior descending coronary artery |
|  | Acute ST segment elevation myocardial infarction due to left coronary artery occlusion |
|  | Acute ST segment elevation myocardial infarction due to right coronary artery occlusion |
| I22.X | Subsequent STEMI of anterior wall |
|  | Subsequent STEMI of interior wall |
|  | Subsequent STEMI of other sites |
|  | Subsequent MI, unspecified |
| Heart Failure (I50) | |
| I50.X | Congestive heart failure |
|  | Left heart failure |
|  | Systolic heart failure |
|  | Diastolic heart failure |
|  | Hypertensive heart failure |
|  | Chronic right-sided heart failure |
|  | Biventricular congestive heart failure |
|  | Acute on chronic right-sided congestive heart failure |
|  | Chronic systolic heart failure |
|  | Chronic diastolic heart failure |
|  | Acute on chronic systolic heart failure |
|  | Acute on chronic diastolic heart failure |
|  | Chronic combined systolic and diastolic heart failure |
|  | Acute on chronic combined systolic and diastolic heart failure |
|  | Acute right-sided heart failure |
|  | Acute systolic heart failure |
|  | Acute diastolic heart failure |
|  | Acute combined systolic and diastolic heart failure |
|  | Heart Failure |
| Peripheral Vascular Diseases (PAD) | |
| I70, 73 | Peripheral vascular disease |
| Composite Cerebrovascular | |
| Stroke (I60-I64) | Subarachnoid hemorrhage |
|  | Intracerebral hemorrhage |
|  | Other nontraumatic intracranial hemorrhage |
|  | Cerebral infarction |
|  | Stroke, not specified as hemorrhage or infraction |
| Transient Ischemic Attack (G45.x) | Transient cerebral ischemic attacks and related syndromes |
| N18.1-N18.6 | Chronic Kidney Disease |
| R80 | Albuminuria |
| I48.X | Afib |
| G47 | Sleep disorder |
| M15-M19 | Osteoarthritis |
| K21 | GERD |
| H40 | Glaucoma |

**Supplementary Table S2.** List of drug names used to identify prescription drug use at baseline and during follow-up

| Drug category | Generic names |
| --- | --- |
| SGLT2 | Canagliflozin  Dapagliflozin  Empagliflozin  Ertugliflozin  Sotagliflozin |
| Sulfonylureas | Glipizide  Glyburide  Glimepiride  Chorpropamide  Tolbutamide  Tolazamide  Acetohexamide |
| DPP4i | Sitagliptin  Saxagliptin  Linagliptin  Alogliptin  Vildagliptin  Teneligliptin |
| Thiazoledinedione | Pioglitazone  Rosiglitazone |
| Insulin | Insulin  Humulin  Humalog  Novolog |
| GLP-1 receptor agonists | Albiglutide  Exenatide  Liraglutide  Lixisenatide  Dulaglutide  Semaglutide  Tirzepatide |
| Other T2D medications | Metformin  Acabose  Miglitol  Repaglinide  Nateglinide  Pramlintide |
| Potential liver benefit medications | Icosapent ethyl  Omega-3-acid ethyl esters  Fenofibrate  Gemfibrozil  Vitamin E  Alpha-tocopherol  Specific statins; atorvastatin, rosuvastatin, pravastatin |
| Anticoagulant | Warfarin  Dabigatran  Rivaroxaban  Apixaban  Edoxaban  Betrixaban  Enoxaparin  Heparin  Fondaparinux |
| Antiplatelets | Anagrelide  Aspirin  Abciximab  Cilostazol  Clopidogrel  Dipyridamole  Eptifibatide  Prasugrel  Icagrelor  Ticlopidine  Vorapaxar  Cangrelor  Tirofiban |
| Statins | Atorvastatin  Simvastatin  Rosuvastatin  Pravastatin  Lovastatin  Fluvastatin  Pitavastatin |
| Other lipid lowering medications | Ezetimibe  Cholestyramine  Colestipol  Colesevelam  Fenofibrate  Gemfibrozil  Niacin  Icosapent ethyl  Omega-3 acid esters |
| PCSK9i | Alirocumab  Evolocumab  Inclisiran |
| MRA | Eplerenone  Spironolactone |
| Non MRA | Finerenone |
| Diuretic | Bendroflumethiazide  Chlorothiazide  Chlorthalidone  Hydrochlorothiazide  Indapamide  Myethoclothiazide  Metolazone  Bumetanide  Ethacrynate sodium  Ethacrynic acid  Furosemide  Torsemide  Amiloride  Triamterene |
| RAAS Inhibitors | Benazepril  Captopril  Enalapril  Fosinopril  Lisinopril  Moexipril  Perindopril  Quinapril  Ramipril  Trandolapril  Azilsartan  Candesartan  Eprosartan  Irbesartan  Losartan  Olmesartan  Telmisartan  Valsartan  Aliskiren |
| Beta Blockers | Atenolol  Betaxolol  Bisoprolol  Metoprolol tartrate  Metoprolol succinate  Nebivolol  Nadolol  Propanolol  Acebutolol  Pindolol  Timolol  Carvedilol  Labetalol  Esmolol  Sotalol |
| Calcium Blockers | Amlodipine  Felodipine  Isradipine  Nicardipine  Nifedipine  Nisoldipine  Clevidipine  Nimodipine  Diltiazem  Verapamil |
| Other antihypertensive medications | Doxazosin  Prazosin  Terazosin  Alfuzosin  Clonidine  Methyldopa  Guanfacine  Hydralazine  Minoxidil  Guenethidine  Tolazoline  Sodium nitroprusside  Phenoxybenzamine hydrochloride  Phentolamine  Fenoldopam |

**Supplementary Table S3.** Baseline characteristics of unmatched cohorts of VINT and Usual Care used for the matching and IPTW analysis

| Characteristics | VINT (N=5,236) | UC (N=200,778) | SMD | p-value |
| --- | --- | --- | --- | --- |
|  | **Mean (SD) or N (%)** | **Mean (SD) or N (%)** |  |  |
| Demographics | | | | |
| Age at registration | 53.0 (9.3) | 46.4 (15.7) | 0.51 | <0.001 |
| Follow-up duration | 805.3 (415.7) | 503.5 (382.4) | 0.76 | <0.001 |
| Gender |  |  | 0.33 | <0.001 |
| Female | 2975 (56.8) | 98456 (49.0) |  |  |
| Male | 2237 (42.7) | 90791 (45.2) |  |  |
| Unknown | 24 (0.5) | 11531 (5.7) |  |  |
| Race/Ethnicity |  |  | 1.01 | <0.001 |
| White | 2807 (64.2) | 39161 (22.9) |  |  |
| Black or African American | 559 (12.8) | 23505 (13.8) |  |  |
| Hispanic or Latino | 404 (9.2) | 25218 (14.8) |  |  |
| Asian or Pacific Islander | 168 (3.8) | 26784 (15.7) |  |  |
| Other | 105 (2.4) | 22490 (13.2) |  |  |
| Unknown | 330 (7.5) | 33718 (19.7) |  |  |
| Baseline comorbidities | | | | |
| Type 2 Diabetes | 2718 (51.9) | 53887 (26.8) | 0.53 | <0.001 |
| Obesity | 1925 (36.8) | 78702 (39.2) | 0.05 | <0.001 |
| MASLD or MASH | 376 (7.2) | 11355 (5.7) | 0.06 | <0.001 |
| MASH | 31 (0.6) | 1002 (0.5) | 0.01 | 0.4 |
| MASLD | 359 (6.9) | 10829 (5.4) | 0.06 | <0.001 |
| Liver procedure | 249 (4.8) | 10858 (5.4) | 0.03 | 0.04 |
| Fibrosis | 11 (0.2) | 248 (0.1) | 0.02 | 0.12 |
| Coronary Artery Disease | 307 (5.9) | 14638 (7.3) | 0.06 | <0.001 |
| Any other cardiovascular disease | 47 (0.9) | 3395 (1.7) | 0.07 | <0.001 |
| Stroke | 88 (1.7) | 7213 (3.6) | 0.12 | <0.001 |
| Atrial Fibrillation | 107 (2.0) | 6335 (3.2) | 0.07 | <0.001 |
| Heart Failure | 68 (1.3) | 7118 (3.5) | 0.15 | <0.001 |
| Hyperlipidemia | 2759 (52.7) | 78645 (39.2) | 0.27 | <0.001 |
| Hypertension | 2705 (51.7) | 80777 (40.2) | 0.23 | <0.001 |
| Chronic Kidney Disease (CKD) | 290 (5.5) | 14470 (7.2) | 0.07 | <0.001 |
| CKD Stage, n (%) |  |  | 0.15 | <0.001 |
| 1 | 17 (0.3) | 443 (0.2) |  |  |
| 2 | 59 (1.1) | 1772 (0.9) |  |  |
| 3 | 28 (0.5) | 1593 (0.8) |  |  |
| 3a | 37 (0.7) | 2250 (1.1) |  |  |
| 3b | 13 (0.2) | 1592 (0.8) |  |  |
| 4 | 2 (0.0) | 789 (0.4) |  |  |
| 5 | 0 (0.0) | 523 (0.3) |  |  |
| Albuminuria | 127 (2.4) | 4195 (2.1) | 0.02 | 0.10 |
| Cancer | 275 (5.3) | 10619 (5.3) | 0.002 | 0.93 |
| Peripheral Artery Disease | 109 (2.1) | 7950 (4.0) | 0.11 | <0.001 |
| Sleep Disorder | 1440 (27.5) | 41474 (20.7) | 0.16 | <0.001 |
| Smoking | 171 (3.3) | 15100 (7.5) | 0.19 | <0.001 |
| Alcohol Use/Abuse | 4359 (2.2) | 49 (0.9) | 0.10 | <0.001 |
| Baseline medications | | | | |
| Potential liver benefit medications or supplements | 2120 (40.5) | 53534 (26.7) | 0.30 | <0.001 |
| Insulin | 486 (9.3) | 11683 (5.8) | 0.13 | <0.001 |
| SGLT2i | 666 (12.7) | 10351 (5.2) | 0.27 | <0.001 |
| Incretin Mimetics | 1086 (20.7) | 20100 (10.0) | 0.30 | <0.001 |
| DPP4i | 313 (6.0) | 5391 (2.7) | 0.16 | <0.001 |
| Sulfonylurea | 276 (5.3) | 4418 (2.2) | 0.16 | <0.001 |
| TZD | 151 (2.9) | 2577 (1.3) | 0.11 | <0.001 |
| Other T2D medications | 2302 (44.0) | 40911 (20.4) | 0.52 | <0.001 |
| Anticoagulant | 140 (2.7) | 8889 (4.4) | 0.10 | <0.001 |
| Antiplatelet | 196 (3.7) | 10387 (5.2) | 0.07 | <0.001 |
| Beta Blocker | 812 (15.5) | 30943 (15.4) | 0.003 | 0.86 |
| Calcium Channel Blocker | 756 (14.4) | 27620 (13.8) | 0.02 | 0.16 |
| Diuretic | 1266 (24.2) | 36234 (18.0) | 0.15 | <0.001 |
| Other Antihypertensive | 132 (2.5) | 9186 (4.6) | 0.11 | <0.001 |
| Statin | 2320 (44.3) | 58722 (29.2) | 0.32 | <0.001 |
| Other Lipid Lowering | 357 (6.8) | 9445 (4.7) | 0.09 | <0.001 |
| PCSK9i | 21 (0.4) | 559 (0.3) | 0.02 | 0.13 |
| Baseline Costs |  |  |  |  |
| Total Cost of Care | 13808.0 (26319.7) | 14903.4 (51055.7) | 0.03 | 0.12 |
| Outpatient Cost | 8206.8 (18488.6) | 8868.9 (42466.8) | 0.02 | 0.26 |
| Inpatient Cost | 1355.0 (9720.7) | 2193.0 (13775.3) | 0.07 | <0.001 |
| Pharmacy Cost | 4233.3 (11521.8) | 3829.4 (17467.6) | 0.03 | 0.10 |
| Baseline area level variables |  |  |  |  |
| Region |  |  | 0.61 | <0.001 |
| Midwest | 2025 (38.7) | 51735 (25.8) |  |  |
| Northeast | 507 (9.7) | 47442 (23.6) |  |  |
| South | 2145 (41.0) | 52512 (26.2) |  |  |
| West | 559 (10.7) | 49083 (24.4) |  |  |
| Area Deprivation Index (ADI) Quintiles | | | 0.34 | <0.001 |
| ADI 1 | 869 (16.6) | 57492 (28.6) |  |  |
| ADI 2 | 2159 (41.2) | 65644 (32.7) |  |  |
| ADI 3 | 1453 (27.8) | 41255 (20.5) |  |  |
| ADI 4 | 483 (9.2) | 21828 (10.9) |  |  |
| ADI 5 | 272 (5.2) | 14581 (7.3) |  |  |

**Note:** Standardized mean differences (SMDs) and *p*-values were calculated using the TableOne package in R. For continuous variables, *p*-values were derived using the student’s *t*-test when the normality assumption was met and the Wilcoxon rank-sum test otherwise. For categorical variables, *p*-values were obtained using the chi-square test or Fisher’s exact test when cell counts were small. SMDs were computed as the absolute difference in means (or proportions for categorical variables) between groups divided by the pooled standard deviation. SMDs were calculated using both the TableOne and MatchIt packages to cross-validate covariate balance estimates. To evaluate covariate balance between matched groups, SMD ≤ 0.1 was used as the criterion for acceptable balance. Unlike *p*-values, which are influenced by sample size and may indicate statistically significant differences that are not clinically meaningful in large samples, SMD provides a sample size–independent measure of balance that quantifies the magnitude of difference between groups. Therefore, SMDs were used as the primary metric for assessing post-matching balance, while *p*-values were reported descriptively.**Abbreviations:** UC, usual care; VINT, Individualized Nutrition Therapy (intervention) program; T2D, type 2 diabetes; MASLD, metabolic dysfunction-associated steatotic liver disease; MASH, metabolic dysfunction-associated steatohepatitis; Afib, atrial fibrillation; PAD, peripheral artery disease; CKD, chronic kidney disease; SGLT2i, sodium–glucose cotransporter-2 inhibitor; DPP4i, dipeptidyl peptidase-4 inhibitor; RAASi, renin–angiotensin–aldosterone system inhibitor (e.g., ACEi/ARB/ARNI); MRA, mineralocorticoid receptor antagonist (steroidal); non-MRA, non-steroidal mineralocorticoid receptor antagonist (e.g., finerenone); PCSK9i, proprotein convertase subtilisin/kexin type 9 inhibitor, potential drugs with liver benefits included Vitamin E, omega 3 fatty acids, fibrates and atorvastatin, rosuvastatin, pravastatin

**Supplementary Table S4.** Comparison of demographics and treatment characteristics between matched VINT participants included in this analysis and unmatched participants or those without closed claims

| Variables | Excluded Cohort, n=40,636 | Matched Final Analytic Cohort, n= 5,031 | SMD |
| --- | --- | --- | --- |
|  | **Mean (SD) or n (%)** | **Mean (SD) or n (%)** |  |
| Age (years) | 53.1 (9.8) | 52.9 (9.3) | 0.01 |
| Gender |  |  | 0.04 |
| Female | 23642 (58.2) | 2853 (56.7) |  |
| Male | 16775 (41.3) | 2161 (43.0) |  |
| Unknown | 219 (0.5) | 17 (0.3) |  |
| Race/Ethnicity |  |  | 0.29 |
| White | 15571 (51.5) | 2682 (64.1) |  |
| African American | 4063 (13.4) | 533 (12.7) |  |
| Hispanic or Latino | 4321 (14.3) | 382 (9.1) |  |
| Asian or Pacific Islander | 1287 (4.3) | 162 (3.9) |  |
| Other | 1058 (3.5) | 98 (2.3) |  |
| Unknown | 3923 (13.0) | 325 (7.8) |  |
| Enrollment BMI Category (%) |  |  | 0.11 |
| <18.5 | 8 (0.0) | 0 (0.0) |  |
| 18.5-24.9 | 1430 (3.5) | 137 (2.7) |  |
| 25.0-29.9 | 6824 (16.8) | 670 (13.3) |  |
| 30.0-34.9 | 12417 (30.6) | 1620 (32.2) |  |
| 35.0-39.9 | 9782 (24.1) | 1276 (25.4) |  |
| ≥40 | 10155 (25.0) | 1327 (26.4) |  |
| Enrollment HbA1c Category (%) |  |  | 0.09 |
| <6.5% | 19165 (49.5) | 2539 (53.8) |  |
| 6.5-<7.0% | 5064 (13.1) | 603 (12.8) |  |
| 7.0-<8.0% | 6559 (17.0) | 693 (14.7) |  |
| 8.0-<9.0% | 3476 (9.0) | 410 (8.7) |  |
| ≥9% | 4419 (11.4) | 472 (10.0) |  |
| Tenure in Virta Program (%) |  |  | 0.04 |
| 12 to 24 months | 23,232 (57.2) | 2980 (59.2) |  |
| ≥ 24 months | 17404 (42.8) | 2051 (40.8) |  |
| Current Virta Program |  |  | 0.18 |
| Diabetes Reversal | 25189 (62.6) | 2816 (56.4) |  |
| Sustainable Weight Loss | 15,447(37.4) | 2215 (43.6) |  |
| 6 Months Mean BHB Category |  |  | 0.06 |
| 0-0.3mM | 10529 (26.5) | 1190 (24.2) |  |
| 0.3-0.5mM | 12132 (30.5) | 1525 (31.0) |  |
| 0.5-1.0mM | 13241 (33.3) | 1700 (34.6) |  |
| ≥ 1mM | 3818 (9.6) | 502 (10.2) |  |
| 6 Months Percentage Weight Change Category |  |  | 0.08 |
| <5% | 13087 (34.0) | 1495 (30.9) |  |
| 5-<10% | 10740 (27.9) | 1352 (27.9) |  |
| 10-<15% | 8320 (21.6) | 1100 (22.7) |  |
| 15-<20% | 4327 (11.2) | 605 (12.5) |  |
| ≥20% | 1998 (5.2) | 286 (5.9) |  |
| 12 Months Percentage Weight Change Category |  |  | 0.06 |
| <5% | 13442 (39.7) | 1524 (37.2) |  |
| 5-<10% | 8017 (23.7) | 1011 (24.7) |  |
| 10-<15% | 5596 (16.5) | 664 (16.2) |  |
| 15-<20% | 3503 (10.3) | 454 (11.1) |  |
| ≥20% | 3311 (9.8) | 442 (10.8) |  |
| 6 Months HbA1c Change Category (%) |  |  | 0.04 |
| <0.5% Decrease | 12079 (47.1) | 1550 (48.9) |  |
| 0.5-1.0% Decrease | 5082 (19.8) | 623 (19.7) |  |
| ≥1.0% Decrease | 8493 (33.1) | 994 (31.4) |  |
| 12 Months HbA1c Change Category (%) |  |  | 0.03 |
| <0.5% Decrease | 11079 (53.0) | 1392 (54.2) |  |
| 0.5-1.0% Decrease | 3796 (18.1) | 463 (18.0) |  |
| ≥1.0% Decrease | 6042 (28.9) | 711 (27.7) |  |
| 6 months diabetes reversal (%) | 15514 (57.4) | 2118 (63.1) | 0.12 |
| 12 months diabetes reversal (%) | 11653 (53.1) | 1656 (61.3) | 0.17 |
| 6 months diabetes remission (%) | 10202 (37.7) | 1410 (42.0) | 0.09 |
| 12 months diabetes remission (%) | 8026 (36.6) | 1176 (43.6) | 0.14 |
| 24 months diabetes remission (%) | 1656 (29.6) | 212 (31.6) | 0.04 |

**Note:** Standardized mean differences (SMDs) and *p*-values were calculated using the TableOne package in R.To evaluate covariate balance between matched groups, SMD ≤ 0.1 was used as the criterion for acceptable balance.

**Abbreviations:** UC, usual care; VINT, Individualized Nutrition Therapy program; BMI, Body mass index; HbA1c, hemoglobin A1c

### **Supplementary Table S5.** Transition Patterns from Baseline to Follow-up Liver Diagnoses among Participants with MASLD, MASH, or Fibrosis at Baseline

| Group | Baseline Diagnosis | Follow-up Diagnosis | n | % of participants with both baseline and follow-up diagnoses |
| --- | --- | --- | --- | --- |
| UC | MASLD | MASLD | 87 | 69.6 |
| UC | Liver_Probable | MASLD | 9 | 7.2 |
| UC | MASLD | Unspecified liver disease | 5 | 4.0 |
| UC | MASLD | Abnormal liver imaging/labs | 4 | 3.2 |
| UC | MASLD | Cirrhosis | 4 | 3.2 |
| UC | MASLD | Hepatic encephalopathy | 4 | 3.2 |
| UC | MASLD | Ascites | 3 | 2.4 |
| UC | Fibrosis | MASLD | 1 | 0.8 |
| UC | Liver_Probable | Abnormal liver imaging/labs | 1 | 0.8 |
| UC | Liver_Probable | Ascites | 1 | 0.8 |
| UC | Liver_Probable | Cirrhosis | 1 | 0.8 |
| UC | Liver_Probable | MASH | 1 | 0.8 |
| UC | Liver_Probable | Unspecified liver disease | 1 | 0.8 |
| UC | Liver_Probable | Varices | 1 | 0.8 |
| UC | MASH | MASH | 1 | 0.8 |
| UC | MASLD | MASH | 1 | 0.8 |
| VINT | MASLD | MASLD | 101 | 74.3 |
| VINT | Liver_Probable | MASLD | 7 | 5.1 |
| VINT | MASLD | Unspecified liver disease | 7 | 5.1 |
| VINT | MASLD | Abnormal liver imaging/labs | 4 | 2.9 |
| VINT | MASLD | Cirrhosis | 4 | 2.9 |
| VINT | Liver_Probable | Unspecified liver disease | 3 | 2.2 |
| VINT | MASLD | Hepatic encephalopathy | 2 | 1.5 |
| VINT | MASLD | Varices | 2 | 1.5 |
| VINT | Fibrosis | MASLD | 1 | 0.7 |
| VINT | Liver_Probable | Abnormal liver imaging/labs | 1 | 0.7 |
| VINT | MASH | MASH | 1 | 0.7 |
| VINT | MASH | Unspecified liver disease | 1 | 0.7 |
| VINT | MASLD | Ascites | 1 | 0.7 |
| VINT | MASLD | MASH | 1 | 0.7 |

**Note:** Transition patterns between baseline and follow-up liver diagnoses are shown for usual care (UC) and VINT participants. Percentages represent the proportion of participants with both baseline and follow-up diagnoses in each treatment group.

**Abbreviations**: UC, usual care; VINT, Virta intervention; MASLD, metabolic dysfunction-associated steatotic liver disease; MASH, metabolic dysfunction-associated steatohepatitis.

**Supplementary Table S6**. Sensitivity Analysis Using Inverse Probability of Treatment Weighting (IPTW): Covariates and Hazard Ratios Across Liver Conditions.

| Covariate | All liver-related diagnoses | | MASH and beyond | | Advanced Liver Disease | | Liver complications | | |
| --- | --- | --- | --- | --- | --- | --- | --- | --- | --- |
|  | HR (95% CI) | p | HR (95% CI) | p | HR (95% CI) | p-value | HR (95% CI) | p-value | |
| VINT | 0.67 (0.51, 0.87) | 0.003 | 0.47 (0.28, 0.78) | 0.004 | 0.40 (0.22, 0.75) | 0.004 | 0.29 (0.14, 0.60) | | <0.001 |
| Age | 1.01 (1.01, 1.02) | <0.001 | 1.03 (1.03, 1.03) | <0.001 | 1.04 (1.03, 1.04) | <0.001 | 1.03 (1.03, 1.04) | | <0.001 |
| Gender | 0.95 (0.91, 1.00) | 0.07 | 0.94 (0.85, 1.04) | 0.22 | 0.94 (0.84, 1.04) | 0.24 | 1.00 (0.89, 1.11) | | 0.95 |
| White | 1.32 (1.20, 1.44) | <0.001 | 1.86 (1.54, 2.24) | <0.001 | 2.05 (1.67, 2.52) | <0.001 | 2.02 (1.63, 2.51) | | <0.001 |
| African American | 1.18 (1.06, 1.31) | 0.00 | 2.25 (1.85, 2.75) | <0.001 | 2.70 (2.18, 3.34) | <0.001 | 2.73 (2.20, 3.40) | | <0.001 |
| Hispanic or Latino | 1.44 (1.30, 1.59) | <0.001 | 1.94 (1.58, 2.38) | <0.001 | 2.07 (1.65, 2.59) | <0.001 | 1.96 (1.55, 2.49) | | <0.001 |
| Other | 1.12 (0.99, 1.28) | 0.07 | 1.14 (0.90, 1.45) | 0.28 | 1.34 (1.03, 1.73) | 0.03 | 1.23 (0.93, 1.61) | | 0.15 |
| Unknown | 0.99 (0.91, 1.09) | 0.88 | 0.93 (0.76, 1.14) | 0.47 | 0.94 (0.75, 1.18) | 0.59 | 0.86 (0.68, 1.09) | | 0.22 |
| Obesity | 1.13 (1.07, 1.19) | <0.001 | 1.07 (0.97, 1.18) | 0.17 | 1.02 (0.92, 1.14) | 0.71 | 1.03 (0.92, 1.15) | | 0.62 |
| Type 2 diabetes | 1.07 (1.00, 1.15) | 0.04 | 1.49 (1.32, 1.67) | <0.001 | 1.55 (1.37, 1.75) | <0.001 | 1.58 (1.39, 1.81) | | <0.001 |
| Incretin Mimetics | 1.24 (1.17, 1.32) | <0.001 | 0.83 (0.74, 0.94) | 0.00 | 0.75 (0.65, 0.86) | <0.001 | 0.68 (0.59, 0.79) | | <0.001 |
| Other potential liver benefit drugs | 1.24 (1.10, 1.40) | <0.001 | 1.18 (0.96, 1.45) | 0.12 | 1.16 (0.94, 1.44) | 0.17 | 1.25 (1.00, 1.58) | | 0.06 |
| SGLT2i | 1.06 (0.98, 1.15) | 0.15 | 0.90 (0.77, 1.05) | 0.20 | 0.92 (0.78, 1.08) | 0.30 | 0.87 (0.73, 1.04) | | 0.13 |
| Statin | 0.99 (0.87, 1.13) | 0.91 | 1.03 (0.83, 1.28) | 0.80 | 1.01 (0.80, 1.27) | 0.93 | 0.98 (0.77, 1.25) | | 0.89 |
| Thiazolidine | 0.87 (0.74, 1.02) | 0.08 | 0.85 (0.63, 1.13) | 0.26 | 0.83 (0.61, 1.13) | 0.24 | 0.84 (0.60, 1.17) | | 0.31 |
| Resmetirom | 4.18 (0.52, 33.53) | 0.18 | nan | nan | nan | nan | nan | | nan |

**Note:** 95% confidence intervals (CIs) were estimated using Cox proportional hazards models weighted by inverse probability of treatment weighting (IPTW). The treatment model included demographic, clinical, and medication covariates at baseline. Separate models were fit for each composite liver outcome: any liver-related diagnosis (“All liver”), MASH and beyond (MASH, fibrosis, cirrhosis, portal hypertension, and hepatocellular carcinoma), and advanced liver disease (fibrosis, cirrhosis, portal hypertension, and hepatocellular carcinoma). IPTW weights were stabilized and trimmed to reduce variance. “VINT” refers to participants enrolled in the Virta Individualized Nutrition Therapy program. Bolded values indicate statistical significance at α = 0.05.

**Supplementary Table S7.** Liver-Specific Imaging and Biopsy Procedures: Counts in the Whole Matched Cohort

| Liver specific imaging and procedures | VINT, N | UC, N |
| --- | --- | --- |
| Biopsy, with imaging | 1 | 1 |
| Biopsy, without imaging | 7 | 18 |
| MRI, abdomen – w & without contrast | 79 | 73 |
| MRI, abdomen – with contrast | 0 | 2 |
| MRI, abdomen – without contrast | 17 | 11 |
| CT scan, abdomen – with contrast | 19 | 18 |
| CT scan, abdomen – w & without contrast | 24 | 17 |
| CT scan, abdomen – without contrast | 16 | 11 |
| Ultrasound, abdomen – complete | 121 | 167 |
| Ultrasound, abdomen – limited | 252 | 275 |
| Liver elastography (FibroScan) | 25 | 25 |
| Paracentesis (diagnostic) | 0 | 0 |
| Paracentesis (therapeutic) | 0 | 10 |
| Endoscopy—variceal surveillance/therapy | 282 | 344 |
| TIPS (creation) | 0 | 1 |
| TIPS (revision) | 0 | 0 |
| Ablation for HCC (laser) | 0 | 0 |
| Ablation for HCC (RF, open) | 0 | 2 |
| Unlisted laparoscopic liver procedure | 2 | 0 |
| Transarterial embolization/chemoembolization (TACE) | 4 | 5 |

**Note:** Combined Wilcoxon rank-sum test for all liver-specific imaging/biopsy indicated a significant difference between groups (p = 0.02).

**Abbreviations:** MRI, magnetic resonance imaging; CT, computed tomography; TIPS, transjugular intrahepatic portosystemic shunt; HCC, hepatocellular carcinoma

**Supplementary Table S8.** Medication Use Patterns in VINT versus Usual Care Groups among baseline and follow-up MASLD/MASH cohorts

| Medication Class | Group | N | Users (n, %) | Mean Days Covered | Median Days | p (use) | p (days) |
| --- | --- | --- | --- | --- | --- | --- | --- |
| Incretin Mimetics | UC | 716 | 306 (42.9%) | 490 | 336 | <0.001 | 0.07 |
|  | VINT | 628 | 327 (52.4%) | 598 | 459 |  |  |
| Potential MASLD Drugs | UC | 716 | 443 (62.0%) | 697 | 540 | 0.18 | 0.10 |
|  | VINT | 628 | 364 (58.3%) | 820 | 660 |  |  |
| SGLT2 Inhibitors | UC | 716 | 138 (19.3%) | 360 | 210 | 0.59 | 0.71 |
|  | VINT | 628 | 129 (20.7%) | 456 | 270 |  |  |
| Statins | UC | 716 | 452 (63.3%) | 673 | 480 | 0.25 | 0.05 |
|  | VINT | 628 | 375 (60.1%) | 782 | 660 |  |  |
| Resmetirom | UC | 716 | 1 (0.1%) | NA | NA | 0.91 | NA |
|  | VINT | 628 | 2 (0.3%) | NA | NA |  |  |

**Note:** Medication classes were defined a priori and included metabolic and liver-directed therapies: GLP-1 receptor agonists (incretin mimetics), SGLT2 inhibitors, thiazolidinediones (pioglitazone; included in potential MASLD drugs), vitamin E, statins, and resmetirom. NA = not applicable due to limited use or follow-up duration.
